# Shared genetic factors between lung function and asthma by age at onset

**DOI:** 10.64898/2026.02.20.26346655

**Authors:** Yining Li, Diana M. Cornejo-Sanchez, Rui Dong, Elnaz Naderi, Gao T. Wang, Suzanne M. Leal, Andrew T. DeWan

**Author notes:** To whom correspondence should be addressed: Dr. Andrew T. DeWan, Department of Chronic Disease Epidemiology, Center for Perinatal, Pediatric and Environmental Epidemiology, Yale School of Public Health, 1 Church Street, 6^th^ Floor, New Haven, Connecticut, 06510, USA.

## Abstract

The genetic relationship between asthma and lung function may be dependent on age-at-onset (AAO) of asthma. We investigated whether the shared genetics between asthma AAO and lung function is dependent on AAO. Asthma cases from UK Biobank were subset according to their AAO and genetic correlation was used to obtain genetically homogeneous groups, i.e., ≤20 (LT20), 20-40, and >40 (GT40) years. Association analysis and fine-mapping were performed to identify shared genetics between AAO groups and lung function. Mediation and quantitative trait locus (QTL) analyses were performed to identify mechanisms underlying shared genetic associations. Chr5, chr6, chr12, and chr17 each had one region that displayed a cross-phenotype replicated association with at least one AAO group and lung function. Overlapping credible sets obtained from fine-mapping were observed on chr5 and chr6. Mediation analyses demonstrated that for each region the proportion mediated through asthma on lung function was larger for asthma LT20 compared to 20-40 and GT40 suggesting that their effects on lung function were more strongly driven by this association. Tissue-specific QTL analysis revealed shared etiology on chr5 may be acting through *SLC22A5* and *C5orf56* which might play an important role in decreased lung function among individuals with earlier-onset asthma.

## Introduction

Asthma is a complex trait characterized by chronic airway inflammation, reversible airflow obstruction, enhanced airway hyperresponsiveness, and bronchospasms induced by a variety of exogenous and endogenous stimuli, including allergens and irritants^1^. It is a heterogeneous disease with different clinical presentations and underlying mechanisms, suggesting a complex etiology involving many different genetic factors^2^. It is the leading chronic disease in children but is also common among adults^3^. Age-at-onset (AAO) impacts asthma heterogeneity. However, the absence of universally accepted and consistently applied AAO categories has substantially constrained efforts to fully characterize the genetic etiology underlying distinct asthma AAO subtypes. Defining asthma AAO groups using genetic correlations could facilitate a more systematic approach to study the genetics of asthma by AAO.

Childhood-onset and adult-onset asthma share genetic risk factors but also have substantial differences in their genetic architecture^4,5^. Genome-wide association studies of childhood-onset and adult-onset asthma have identified many important genes, e.g., *IL1RL1*, *SMAD3*, *FLG* as well as the 17q12-21 locus that includes *ORMDL3*, *GSDMA*, and *GSDMB* which are strongly associated with childhood but not adult asthma^2,4–6^.

Previous studies have shown that the development of lung function abnormalities is a complicated process and involves many shared mechanisms with asthma etiology, such as bronchospasm of airway smooth muscle and inflammation of the airway walls^7^. Genetic factors play a critical role in determining lung function. The most extensive meta-analysis of lung function (N∼590,000 individuals) has identified >1,000 independent signals including *SMAD3* and *TGFB2*^8,9^.

A common clinical test to measure lung function is spirometry with the key metrics being forced expiratory volume in 1 second (FEV1), forced vital capacity (FVC), as well as their ratio (FEV1/FVC). According to the Global Initiative for Asthma Main Report 2024^10^, lung function should be assessed at diagnosis and regularly during asthma treatment since its evaluation can help with treatment response assessment, asthma exacerbation identification, and trigger evaluation^10^.

Few studies have focused on the shared genetic architecture between asthma and lung function and there is a limited understanding of their shared genetics. We used a novel approach to delineate asthma AAO groups using genetic correlations followed by genome-wide association studies (GWAS) using large biobank data to determine pleiotropic loci for asthma and lung function. Mediation analyses of pleiotropic loci demonstrated that for each region the effect on lung function was more strongly driven by earlier onset asthma compared to when onset was >20 years-of-age.

## Methods

### Data access and ethical approval

This research was conducted using data obtained from the UK Biobank Resource (application number 32285). The UK Biobank study received generic approval from the National Research Ethics Service of the National Health Services. The analyses presented in this study were approved by the Human Investigations Committee at Yale University (2000026836) and the Institutional Review Board at Columbia University (IRB-AAAS3494).

### Discovery dataset

Genotyping was performed for 488,282 UK Biobank study subjects using either the UK BiLEVE array or the UK Biobank Axiom array, with 733,332 autosomal variants common to both. Details of subject- and variant-level quality control were previously described.^11^ Analyses were restricted to participants who self-identified as White British (f.22006) and demonstrated similar genetic ancestry based on principal component (PC) analysis (N = 366,752 study subjects)^11^. Variants included in the analysis met the following criteria: call rate >99%, did not deviate from Hardy-Weinberg equilibrium (p-value >5 × 10_J_J), and had a minor allele frequency (MAF) >0.01 (N = 529,024). These genotype array variants were used to generate PCs, perform ridge regression as implemented in REGENIE^12^, compute summary statistics and LD scores for genetic correlations, and estimate heritabilities. Imputed variants provided by the UK Biobank that had a MAF >0.001 and Info Score >0.8 (N = 13,407,279) were used for association and mediation analysis.

For analyses requiring unrelated individuals, we used those study subjects more distantly related than third degree relatives (f.22020). There were 307,259 unrelated White British study subjects for downstream analysis^11^.

### Replication dataset

For the replication cohort, we identified a subset of non-British White European individuals based on self-reported ancestry (f.21000) and PC analysis (f.22009). Specifically, we calculated the mean and covariance for the first 40 PCs among genetically confirmed White British individuals. Mahalanobis distances from this empirical PC distribution were then computed for all participants. Using a Mahalanobis distance threshold that captures 94% (N=458,967) of subjects, we capture all genetically confirmed White British study subjects, 46,589 self-reported White European non-British study subjects, and 2,763 who did not self-report of being of White ethnicity (e.g. “White and Black Caribbean”, “White and Black African”, “White and Asian”, “Any other mixed background”, “Other ethnic group”, “Do not know”, “Prefer not to answer”). The 49,352 White European non-British subjects were then moved forward to the replication dataset. Following quality control, the final replication dataset comprised 44,173 non-British White European individuals^11^, with 13,243,888 imputed and 550,028 directly genotyped variants available for analysis.

### Phenotype definitions

Asthma was defined using either ICD-10 code (f.41270, J45 or J46) or diagnosis by a doctor which was self-reported by the study subject (f.6152) at the initial assessment visit. Age at diagnosis of asthma was self-reported (f.3786). Controls had no report of asthma or autoimmune disease [self-report of a doctor’s diagnosis of autoimmune disease^13^ (f.20002) or sarcoidosis (f.22133)]. Participants who did not report asthma at recruitment but reported it at a subsequent visit were excluded from both asthma cases and controls. Lung function was defined as a continuous variable using the ratio of the best measure for FEV1 (f.20150) to the best measure for FVC (f.20151) which both were the result of an acceptable lung function test (f.3061). In the discovery dataset, there were 48,623 cases and 290,722 controls for asthma and 277,129 study subjects with lung function. In the replication sample, there were 5,822 cases and 36,615 controls for asthma and 30,724 study subjects with lung function.

### Phenotype specific principal components analyses

Linkage disequilibrium (LD) pruning was applied to the directly genotyped variants using the default parameters of PLINK2.0^14,15^ (r^2^ < 0.2). PC analysis was performed independently for the study subjects included in the analysis of each trait using smartPCA in EIGENSOFT-7.2.1^16^. The first 10 PCs were included in the univariate analyses.

### Asthma AAO grouping and genetic correlation (r_g_)

To obtain genetically homogeneous groups based on AAO, an agnostic approach was used (**Figure 1**). Association analysis was performed for the discovery sample using REGENIE^12^ which approximates a linear mixed model (LMM) or generalized LMM (GLMM) by applying ridge regression. Using genotype variant data, association analysis was performed for individuals with asthma onset in 10-year intervals, i.e., ≤10, 10-20, 20-30, 30-40, >40. For each age interval the same set of controls were used in the analysis. Included as covariates in the analyses were age-at-recruitment (f.21022), genetic sex (f.22001), and the first 10 PCs. Summary statistics from each 10-year AAO window were used to compute pairwise r_g_ using LD score regression (LDSC)^17^. We used in-sample LD scores calculated from the same genotyped variants within all asthma cases and a random 1:1 sample of controls. We combined AAO groups iteratively starting with the pair that had the highest r_g_ >0.8 and then recalculated the pairwise r_g_. We continued this process until the remaining r_g_ were <0.8. The resulting three AAO groups were asthma LT20, 20-40, and GT40. Additionally, association analysis was performed for lung function (FEV1/FVC) including the same covariates as were used for asthma and also baseline smoking status (f.20116: never, previous, or current tobacco smoking). Genotype array data from the discovery sample was used to obtain summary statistics for the analyses. LDSC was then used to obtained genetic correlations between asthma LT20, 20-40, GT40, and lung function.

**Figure 1.**
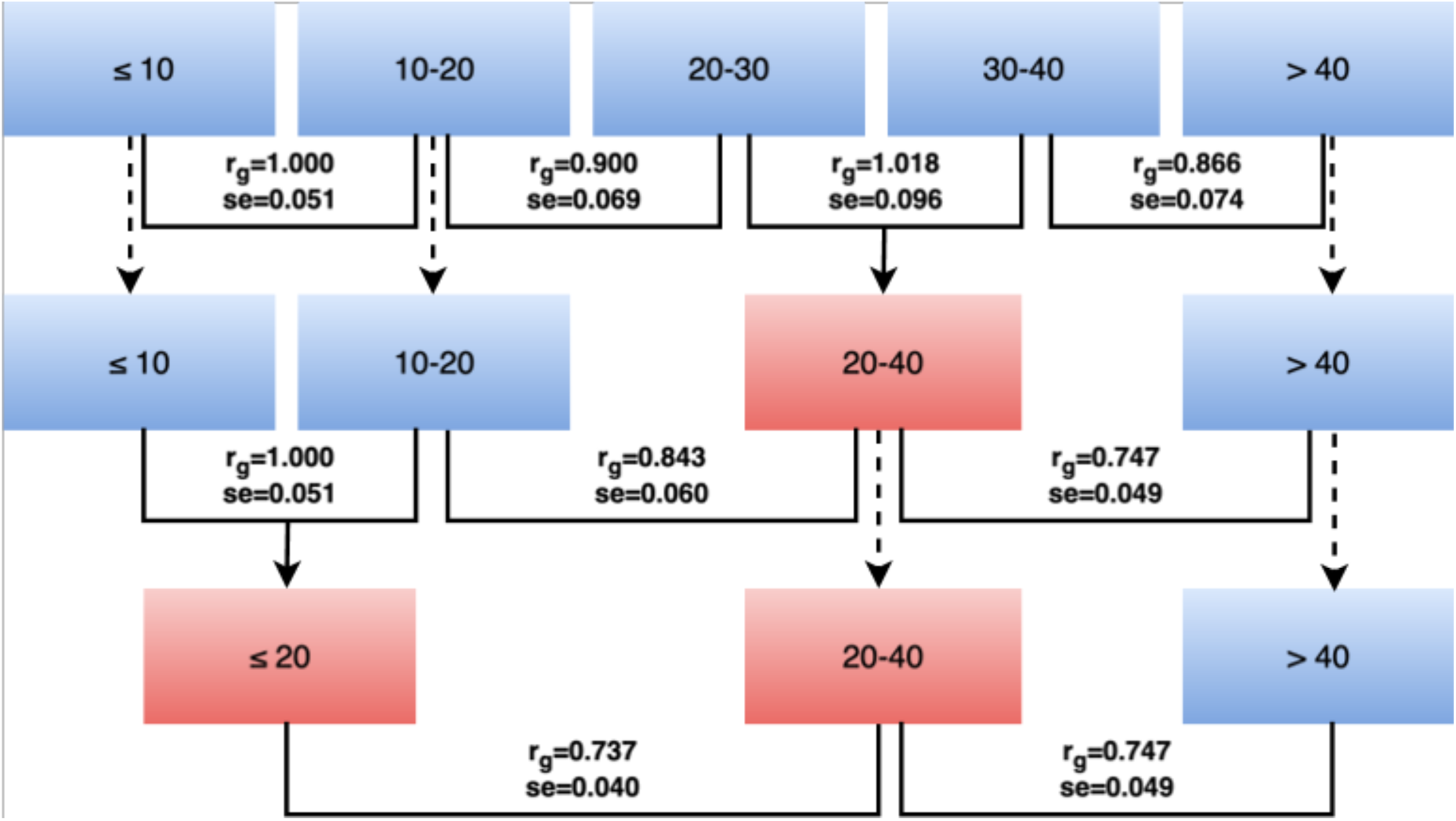
Stepwise clustering of asthma age-at-onset (AAO) using pairwise genetic correlations. Pairwise genetic correlations were calculated using LD score regression (LDSC) starting with 10-year AAO groups (≤10, 10-20, 20-30, 30-40, >40) in the discovery dataset combining the two groups with the highest correlation in each step until all the pairwise genetic correlations were <0.8. In-sample LD scores were estimated from genotype array variants using all asthma cases and a random 1:1 sample of controls. Abbreviations: r_g_: genetic correlation. se: standard error.

### Heritability estimates

To estimate the narrow-sense heritability for each of the asthma AAO groups and lung function, we employed genomic relatedness matrix (GRM) restricted maximum likelihood (GREML) in genome-wide complex trait analysis (GCTA)^18–20^ using genotype array data from the discovery sample. For the liability scale, asthma prevalence for each AAO was estimated from UK Biobank.

### Univariate analyses

Association analysis was performed separately for discovery and replication samples using REGENIE^12^ for LT20, 20-40, GT40 and lung function using the imputed variant data. The same covariates were used as those used to obtain summary statistics for the genetic correlations. Variants in the discovery sample with a p-value<5x10^-8^ were considered to be statistically significant.

### Detection of potentially pleiotropic loci

Variants within a region that were statistically significant in the discovery sample for more than one phenotype were selected. To highlight the role of variants outside of the HLA region, this region was excluded from pleiotropy analysis.

### Detection of lead variants

FUMA 1.6.1^21^ was used to annotate variants to genes. The default r^2^ thresholds were applied to define the border of the risk locus (r^2^ ≥ 0.6) and the lead variant within the locus (r^2^ ≥ 0.1). 1000 Genomes phase 3 European data was used to construct the LD reference panel. Summary statistics were used to determine the lead variant for each phenotype with at least one significant variant in the region.

### Region selection and replication

LocusZoom^22^ was used to generate regional plots based on summary statistics. In the discovery sample, a region was selected if it contained one or more significant variants between at least one asthma AAO group and lung function. The region was considered replicated if the discovery sample lead variant had a replication p-value < 0.05 with a beta in the same direction. Selected regions were merged if they had positional overlaps.

### Fine-mapping

To identify a set of causal independent variants for the overlapping replicated regions, the Sum of Single Effects (SuSiE) model^23,24^ was used. In-sample LD matrices were estimated using the discovery sample. Fine-mapping was performed using summary statistics assuming a maximum of 10 casual variants with non-zero effects to generate a 95% credible set (CS) for each region. Additionally, multivariate SuSiE (mvSuSiE)^25^ was applied to improve power and comprehensively identify shared causal variants across phenotypes. mvSuSiE also used a maximum of 10 casual variants which in the prior were allocated equal weights and a residual variance matrix was used to account for correlations across the phenotypes. For each cross-trait CS, a conditional local false sign rate (clfsr) < 0.01 was used as the threshold for each variant in each trait^25^. The variant was the largest posterior inclusion probability (PIP) in each CS with a clfsr < 0.01 for at least one asthma AAO group and lung function was selected for mediation analysis.

### Mediation analysis

To dissect cross-phenotype associations and decompose the effect of variants on lung function both directly and indirectly through asthma, the R package *mediation* was used^26^ including the variants obtained from fine-mapping. Asthma AAO groups were used as the mediator considering the temporality between asthma and lung function measurements. Given that *mediation* can only use standard linear or logistic regression, only unrelated individuals (N=307,259) were analyzed. For the quasi-Bayesian approximation, 1,000 Monte Carlo draws were used ^26^. A significance threshold of α<0.05 was used to evaluate the total, direct, and indirect effects.

### Expression/splicing quantitative trait locus (eQTL/sQTL) analysis

For the same variants analyzed in the mediation analysis, it was investigated if they were a *cis* eQTLs/sQTLs in relevant tissues, i.e., lung, spleen, whole blood, fibroblasts, skin, and lymphocytes^3^ using the European subset of Genotype-Tissue Expression (GTEx) Project Analysis Release V8 (dbGaP Accession phs000424.v8.p2)^27,28^. Significant QTLs were selected when the nominal p-value was less than the gene-specific p-value threshold which was determined using permutation testing^27,28^. Cis-variants were defined as those residing within 1Mb up- or down-stream of the transcriptional start site.

## Results

Three asthma AAO groups were obtained from genetic correlation analysis: LT20, 20-40, and GT40 (**Figure 1**). The genetic correlations were 0.737 between asthma LT20 and 20-40 (p-value=1.53x10^-75^) and 0.747 between 20-40 and GT40 (p-value=2.15x10^-52^) (**Tables 2** and **S1a-c**). Sample sizes for each asthma AAO group as well as baseline characteristics are displayed in **Table 1**. Heritability was estimated for LT20 (0.397), 20-40 (0.181), GT40 (0.156), and lung function (0.248) (**Tables 2** and **S1d**).

**Table 1.**
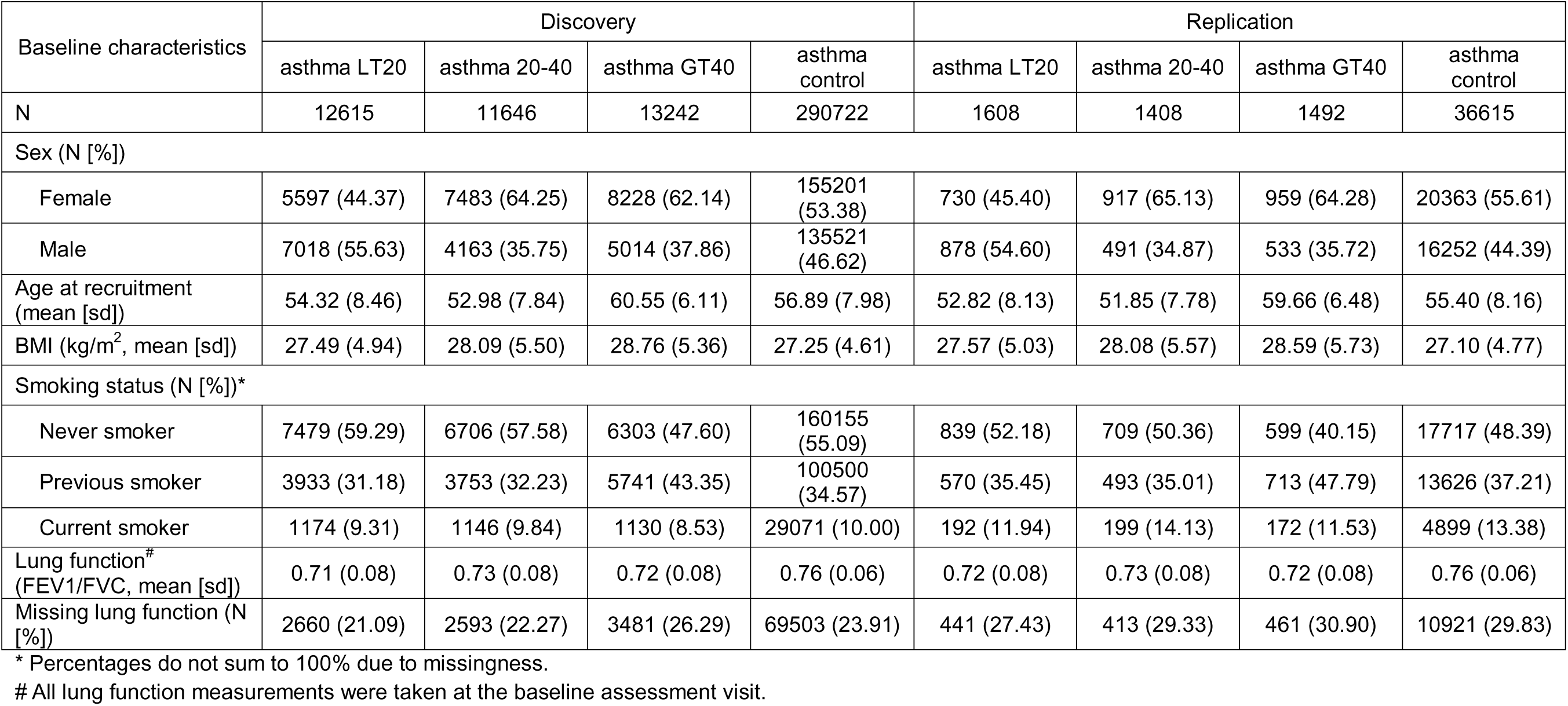
Discovery and replication sample demographics.

| Baseline characteristics | Discovery |  |  |  | Replication |  |  |  |
| --- | --- | --- | --- | --- | --- | --- | --- | --- |
|  | asthma LT20 | asthma 20-40 | asthma GT40 | asthma control | asthma LT20 | asthma 20-40 | asthma GT40 | asthma control |
| N | 12615 | 11646 | 13242 | 290722 | 1608 | 1408 | 1492 | 36615 |
| Sex (N [%]) |  |  |  |  |  |  |  |  |
| Female | 5597 (44.37) | 7483 (64.25) | 8228 (62.14) | 155201 (53.38) | 730 (45.40) | 917 (65.13) | 959 (64.28) | 20363 (55.61) |
| Male | 7018 (55.63) | 4163 (35.75) | 5014 (37.86) | 135521 (46.62) | 878 (54.60) | 491 (34.87) | 533 (35.72) | 16252 (44.39) |
| Age at recruitment (mean [sd]) | 54.32 (8.46) | 52.98 (7.84) | 60.55 (6.11) | 56.89 (7.98) | 52.82 (8.13) | 51.85 (7.78) | 59.66 (6.48) | 55.40 (8.16) |
| BMI (kg/m<sup>2</sup>, mean [sd]) | 27.49 (4.94) | 28.09 (5.50) | 28.76 (5.36) | 27.25 (4.61) | 27.57 (5.03) | 28.08 (5.57) | 28.59 (5.73) | 27.10 (4.77) |
| Smoking status (N [%])* |  |  |  |  |  |  |  |  |
| Never smoker | 7479 (59.29) | 6706 (57.58) | 6303 (47.60) | 160155 (55.09) | 839 (52.18) | 709 (50.36) | 599 (40.15) | 17717 (48.39) |
| Previous smoker | 3933 (31.18) | 3753 (32.23) | 5741 (43.35) | 100500 (34.57) | 570 (35.45) | 493 (35.01) | 713 (47.79) | 13626 (37.21) |
| Current smoker | 1174 (9.31) | 1146 (9.84) | 1130 (8.53) | 29071 (10.00) | 192 (11.94) | 199 (14.13) | 172 (11.53) | 4899 (13.38) |
| Lung function<sup>#</sup> (FEV1/FVC, mean [sd]) | 0.71 (0.08) | 0.73 (0.08) | 0.72 (0.08) | 0.76 (0.06) | 0.72 (0.08) | 0.73 (0.08) | 0.72 (0.08) | 0.76 (0.06) |
| Missing lung function (N [%]) | 2660 (21.09) | 2593 (22.27) | 3481 (26.29) | 69503 (23.91) | 441 (27.43) | 413 (29.33) | 461 (30.90) | 10921 (29.83) |

\* Percentages do not sum to 100% due to missingness.

**Table 2.**
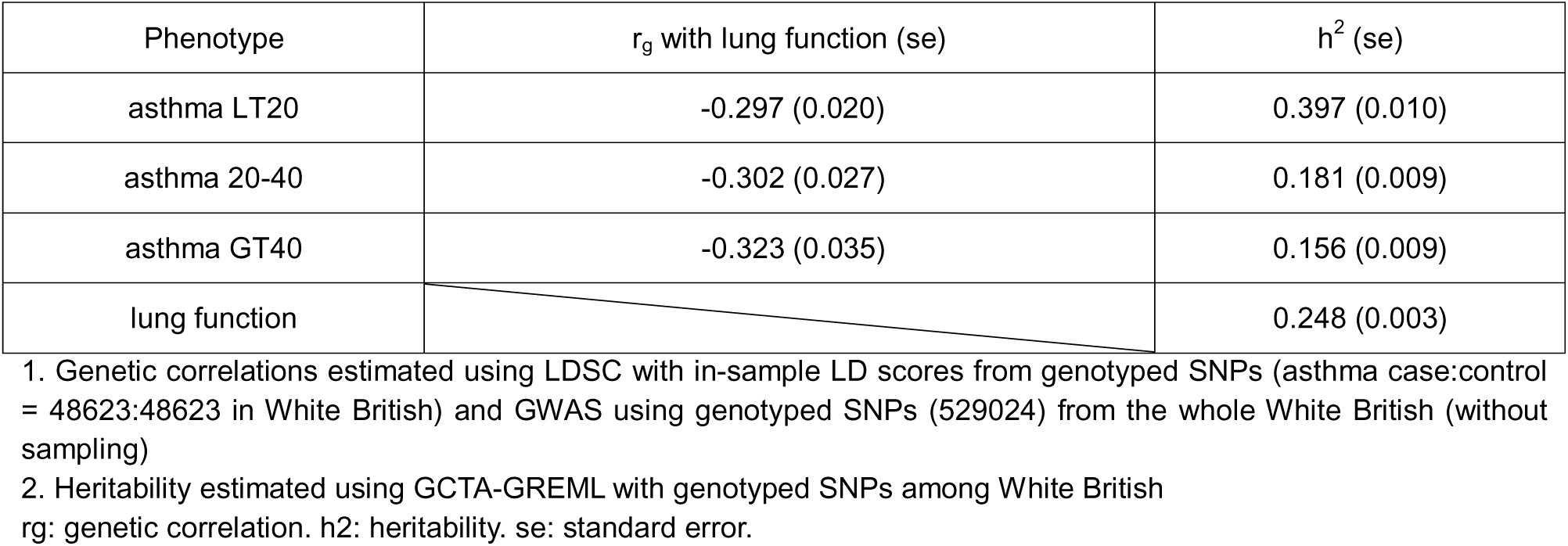
Genetic correlations between asthma AAO subtypes and lung function with heritability estimates.

The number of significant variants for each phenotype are shown in **Figure S1** There were 19 variants significant for all three asthma AAO groups and lung function (**Figure S1)**. However, none of the 19 shared variants replicated across all phenotypes (**Table S2a**), although there were a few variants replicated for at least one asthma AAO group and lung function.

The number of genes with at least one significantly associated variant was determined for LT20 (N=583), 20-40 (N=71), GT40 (N=20), and lung function (N=2,053). Nine genes were shared across the three AAO groups and lung function (**Figure 2, Table S3**). Example LocusZoom plots are shown in **Figure S3** for one region that was significant and replicated for asthma LT20 and lung function.

**Figure 2.**
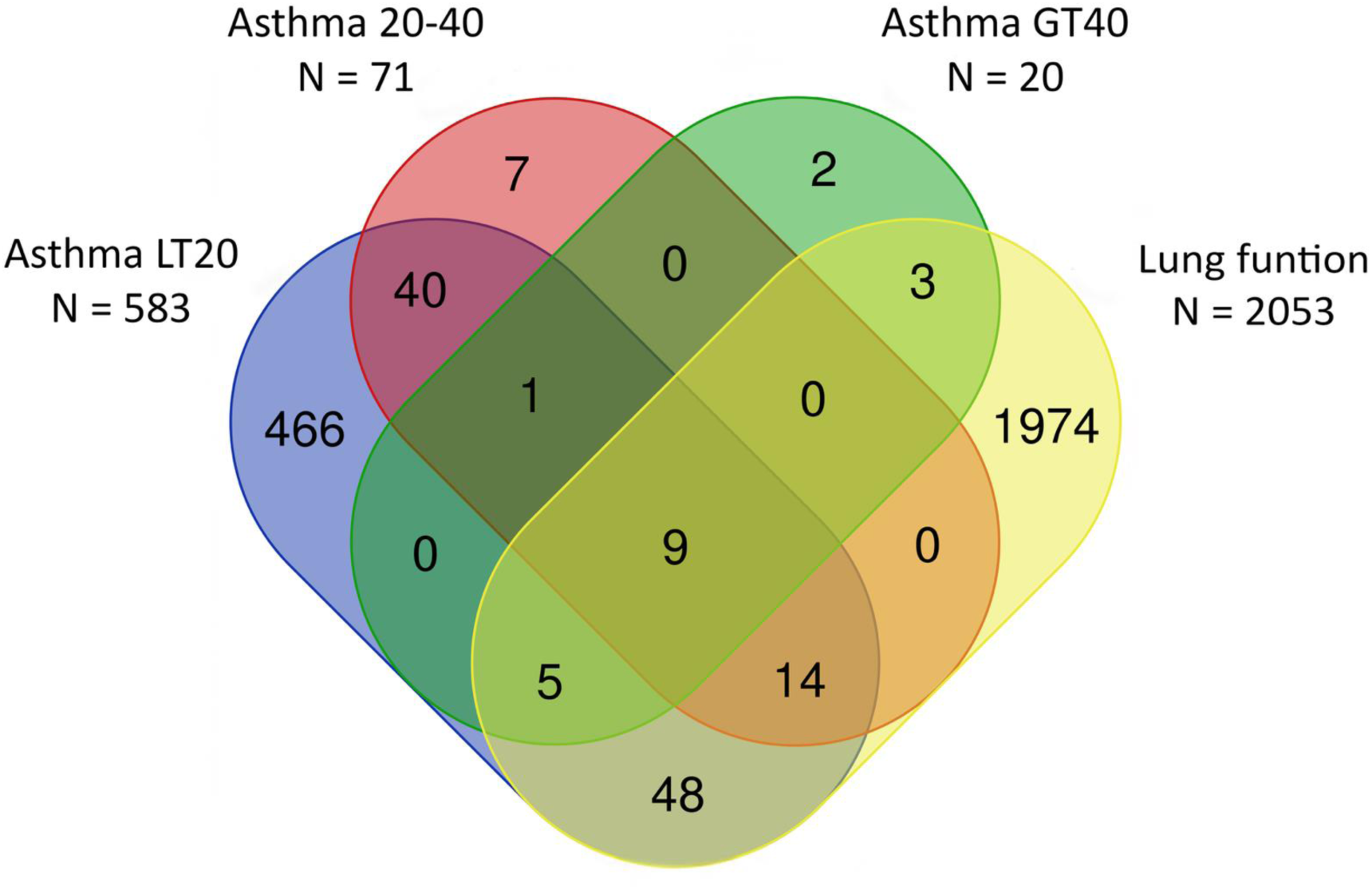
Venn diagram of overlapping genes between asthma age-at-onset groups and lung function. The total number of genes for each trait is provided below their name.

Eighteen regions were carried forward for replication (**Table S4**), and seven regions were replicated in at least one asthma AAO group and lung function which consisted of four independent regions after merging for which fine-mapping was performed (**Table 3**, **Figure S2**).

**Table 3.** Summary of replicated regions.

| Before merging |  |  |  |
| --- | --- | --- | --- |
| Chromosome | Position in hg19 | Group | Phenotype replicated |
| 5 | 131640000-131834000 | asthma LT20, asthma 20-40, asthma GT40, lung function | asthma LT20, asthma GT40, lung function |
| 5 | 131402400-131834000 | asthma LT20, asthma GT40, lung function | asthma LT20, asthma GT40, lung function |
| 5 | 131402400-131834000 | asthma GT40, lung function | asthma GT40, lung function |
| 6 | 90827100-91020000 | asthma LT20, asthma 20-40, lung function | asthma LT20, lung function |
| 12 | 57489000-57578000 | asthma LT20, asthma 20-40, lung function | asthma LT20, asthma 20-40, lung function |
| 12 | 57489000-57578000 | asthma LT20, lung function | asthma LT20, lung function |
| 17 | 38103000-38410000 | asthma LT20, lung function | asthma LT20, lung function |
| After merging |  |  |  |
| Chromosome | Position in hg19 | Group | Phenotype replicated |
| 5 | 131402400-131834000 | asthma LT20, asthma 20-40, asthma GT40, lung function | asthma LT20, asthma GT40, lung function |
| 6 | 90827100-91020000 | asthma LT20, asthma 20-40, lung function | asthma LT20, lung function |
| 12 | 57489000-57578000 | asthma LT20, asthma 20-40, lung function | asthma LT20, asthma 20-40, lung function |
| 17 | 38103000-38410000 | asthma LT20, lung function | asthma LT20, lung function |

For chr5, one CS was identified in both univariate and multivariate fine-mapping. Although univariate fine-mapping did not support colocalization across all four phenotypes, multivariate fine-mapping identified 4 shared variants for each of the four traits (**Figures 3A and S4**). All 4 variants were in the intronic region of *C5orf56* (**Table S5a**). Variant rs7713065, which had the highest PIP of the four, was selected for mediation analysis. There was a strong direct effect in all analyses as well as a smaller but significant indirect effect. In addition, the total effect of the variant on lung function as well as the proportion mediated decreased with increasing asthma AAO (**Table 4**). For the QTL analysis in lung tissue, rs7713065 was a *cis* eQTL for *SLC22A5* and a *cis* sQTL for *P4HA2* and *C5orf56* (**Table S6a-b**). Results for additional tissues can also be found in **Table S6a-b**.

**Figure 3.**
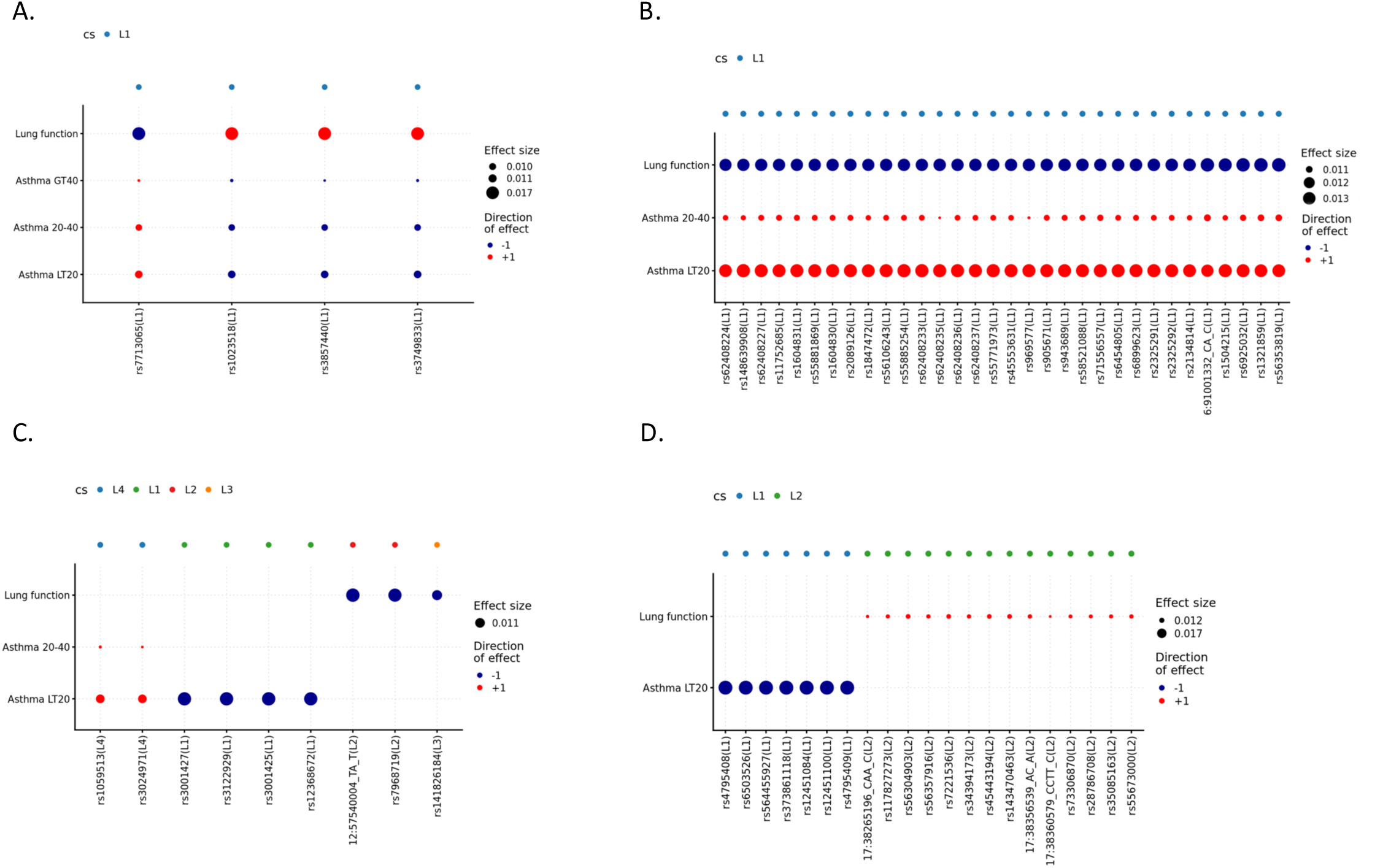
Results from multivariate fine-mapping. The figure displays results for chromosomes 5 (A), 6 (B), 12 (C) and 17 (D). Each credible set (CS) is labelled with L#.

**Table 4.**
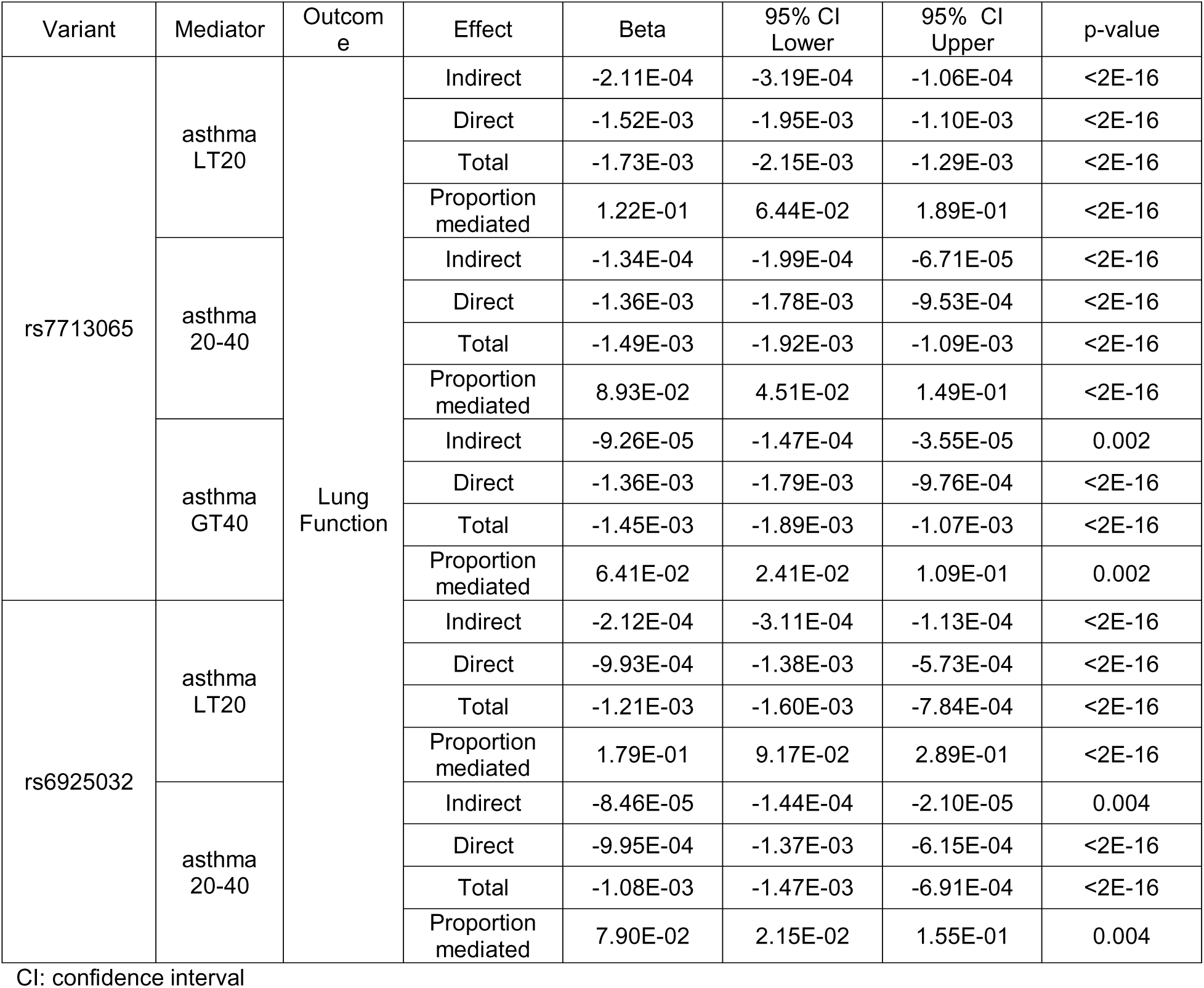
Mediation analyses of top variants in the chromosome 5 and 6 regions.

One CS was identified for the univariate and multivariate fine-mapping on chr6 (**Figure S5**). Both analyses showed strong evidence of colocalization across LT20, 20-40, and lung function (**Figure 3B**). The majority of variants were in the intronic region of *BACH2*, and three variants had *BACH2* as the nearest upstream gene (**Table S5b**). For the variant with the highest PIP, rs6925032, a similar trend of decreasing total effect and proportion mediated by asthma on lung function with increasing asthma AAO was observed (**Table 4**). However, rs6925032 was only a *cis* eQTL for *BACH2* in the pancreas tissue (**Table S6a**). Results for additional tissues can be found in **Table S6a-b**.

For the region on chr12, univariate fine-mapping identified two CSs for LT20, one CS for 20-40, and two CSs for lung function (**Figure S6**). Multivariate fine-mapping revealed four CSs in the chr12 region (**Figure S6**). Neither univariate nor multivariate fine-mapping showed evidence of colocalization between LT20, 20-40, and lung function (**Figure 3C, Table S5c**) therefore no variants were selected for mediation analysis.

Univariate fine-mapping of the chr17 region identified four CSs for LT20 and one for lung function which did not overlap. For multivariate fine-mapping, two non-overlapping CSs were identified one for each phenotype (**Figure 3D**, **Figure S7**). The univariate and multivariate fine-mapping results were consistent with no evidence of colocalization across the region (**Table S5d**).

## Discussion

One aspect that contributes to asthma heterogeneity is AAO. Previous studies have shown that there are distinct genetic associations based on AAO^4,5,29–31^; however, there are no clear age strata. Therefore, genetic correlation analysis was used to define genetically homogenous subtypes of asthma.

The three asthma AAO groups showed similar genetic correlations with lung function. Comparisons of significant results in the discovery sample, showed that shared single variant and gene-region signals between asthma and lung function decreased with increasing asthma AAO. As the sample sizes were similar for asthma AAO groups, this decrease was unlikely a result of lower power. Genetic correlation summarizes the average effect of pleiotropy across all causal loci^32^, and approximately the same genetic correlations do not necessarily indicate a similar number of shared genes or variants between phenotypes. Additionally, there was a decrease in heritability with increasing asthma AAO while genetic correlations remained similar with lung function. This may suggest that later AAO and lung function share a larger number of weaker genetic signals compared to earlier AAO or could be an artifact of the methods used to estimate heritability and genetic correlation.

Univariate and/or multivariate fine-mapping demonstrated that two of the regions overlapped between at least one asthma AAO group and lung function. Mediation analysis suggests that the chr5 and chr6 associations with lung function are more strongly driven by their associations with earlier onset asthma. These regions were investigated in the downstream analyses.

eQTL mapping using rs7713065, which had the highest PIP in the chr5 CS, pointed to genes in relevant tissues, including *AC116366.6* (blood), *PDLIM4* (skin), *and SLC22A5* [blood, lung, fibroblasts, skin, skin (not exposed to sun), spleen]. Additionally, sQTL mapping identified the following genes *C5orf56* (blood, lung), *IRF1* (blood, lung), *P4HA2* [lung, skin, skin (not exposed to sun), and spleen], and *SLC22A4* (blood).

In previous studies, *SLC22A5*, *SLC22A4*, and *P4HA2* mainly showed association with asthma (ever), white blood cell counts, and metabolism-related traits^29,33–45^ and *P4HA2* is also associated with lung function^8,45^. *PDLIM4* is associated with lung function, metabolism-related traits, and inflammatory bowel disease^46–48^. In addition to the traits mentioned, *IRF1* is also associated with childhood- and adult-onset asthma^4,5,49^, asthma AAO^50^, chronic obstructive pulmonary disease (COPD)^51^, and a composite phenotype of asthma and COPD^52^. *C5orf56* is associated with this composite phenotype as well^52^. *SLC22A5* and *SLC22A4* are part of the solute carrier family 2. The encoded protein of *SLC22A5* is an important organic cation transporter and sodium-dependent high affinity carnitine transporter that plays a central role in fatty acid metabolism and energy production within cells^53–58^. *SLC22A4* also encodes an organic cation transporter that is involved in transmembrane ergothioeine^59^ and acetylcholine transportation^60^, and potentially plays a role in controlling inflammation and oxidative stress^60,61^. *P4HA2* encodes a subunit of prolyl 4-hydroxylase, a key enzyme involved in collagen biosynthesis. *PDLIM4* is a gene that encodes a protein involved in cytoskeletal organization and cell signaling^62^. *IRF1* is a member in the interferon regulatory factor gene family and *IRF1-AS1* is the long non-coding RNA regulating its expression and function. Together, they regulate host response to viral and bacterial infections as well as genes expressed in many inflammation and immune processes^63–70^.

The chr6 region showed consistent evidence of overlap for both univariate and multivariate fine-mapping for LT20, 20-40, and lung function with rs6925032 having the highest PIP. *BACH2* was the only gene for which rs6925032 is an eQTL and no sQTLs were identified. Previous studies showed that *BACH2* is associated with various immune-related diseases, including asthma (childhood-onset^4,5,43,49^ and adult-onset^4,5,49^) and asthma AAO^4,50^, blood cells^37,38,42–45^, FEV1/FVC^8,45^, and COPD^51^. *BACH2* is a basic region-leucine zipper domain transcription factor. It regulates transcriptional activity during the differentiation and maturation of T- and B-lymphocytes^71^ and plays an important role in T helper cell 2 (Th2)-mediated inflammatory diseases^72^.

The results suggest that genetics plays a greater role in the relationship between earlier-onset asthma and lung function with QTL mapping suggesting that immune-mediated processes are the main factors underlying this relationship. However, non-genetic factors (e.g., air pollution) might have a stronger effect in the shared etiology between later-onset asthma and lung function.

Our study had several limitations. First, asthma was defined based on self-report of a doctor’s diagnosis and electronic health records. It is possible that the diagnosis of asthma or asthma AAO was mis-specified for some participants. Our sample of adult asthmatics may also include individuals with early-onset asthma whose symptoms resolved but reappeared later. Second, the UK Biobank cohort is prone to a “healthy volunteer” selection bias, where the study population is not representative of the target population and tends to be healthier and of higher socio-economic status^73^. Third, our analyses focused on individuals of European ancestry due to the composition of the UK Biobank. Finally, our QTL analysis was limited to *cis* eQTLs/sQTLs, but given the relatively small sample size of GTEx, *trans* QTLs were unable to be studied due to their weak effects.

The agnostic grouping of asthma by AAO using genetic correlations is a novel approach that can also be used for other complex traits with broad AAO distributions, e.g., type 2 diabetes, testicular cancer, to create more genetically homogeneous groups. Genetic analysis of asthma informed by AAO may assist in explaining differences in pathophysiology. Using asthma AAO groups allowed for the dissection of the genetic relationship between asthma and lung function and pointing to the most critical AAO group for the genetic associations for asthma impacting lung function.

## Gene Abbreviations

BACH2: BTB domain and CNC homolog 2
C5orf56: Chromosome 5 open reading frame 56, which is also known as *CARINH* (colitis associated IRF1 antisense regulator of intestinal homeostasis) and *IRF-AS1* (IRF1 antisense RNA 1)
FLG: Filaggrin
GSDMA: Gasdermin A
GSDMB: Gasdermin B
IL1RL1: Interleukin 1 receptor-like 1
IRF1: Interferon regulatory factor 1
ORMDL3: ORMDL sphingolipid biosynthesis regulator 3
P4HA2: Prolyl 4-hydroxylase subunit alpha 2
PDLIM4: PDZ And LIM domain 4
SLC22A4: Solute carrier family 22 member 4
SLC22A5: Solute carrier family 22 member 5
SMAD3: Mothers against decapentaplegic homolog 3
TGFB2: Transforming growth factor beta 2

## Acknowledgements

The authors would like to thank the researchers and participants of the United Kingdom Biobank. All data was accessed as part of project 32285 from the United Kingdom Biobank. We thank the Yale Center for Research Computing for the use of the McCleary High Performance Computing cluster.

## Data Availability

UK Biobank individual-level genotype and phenotype data, from which were used in this study are available to approved researchers. Instructions for access to UK Biobank data are available at https://www.ukbiobank.ac.uk/enable-your-research. Summary statistics for each GWAS are all available in the GWAS Catalog (GCST90809793, GCST90809794, GCST90809795, GCST90809796, GCST90809797, GCST90809798, GCST90809799, GCST90809800).

## References

1. Tomisa G, Horváth A, Sánta B, Keglevich A, Tamási L. Epidemiology of comorbidities and their association with asthma control. Allergy Asthma Clin Immunol. 2021;17(1):95. doi:10.1186/s13223-021-00598-3

2. Papi A, Brightling C, Pedersen SE, Reddel HK. Asthma. The Lancet. 2018;391(10122):783-800. doi:10.1016/S0140-6736(17)33311-1

3. El-Husseini ZW, Gosens R, Dekker F, Koppelman GH. The genetics of asthma and the promise of genomics-guided drug target discovery. Lancet Respir Med. 2020;8(10):1045–1056. doi:10.1016/S2213-2600(20)30363-5

4. Ferreira MAR, Mathur R, Vonk JM, et al. Genetic Architectures of Childhood- and Adult-Onset Asthma Are Partly Distinct. Am J Hum Genet. 2019;104(4):665–684. doi:10.1016/j.ajhg.2019.02.022

5. Pividori M, Schoettler N, Nicolae DL, Ober C, Im HK. Shared and distinct genetic risk factors for childhood-onset and adult-onset asthma: genome-wide and transcriptome-wide studies. Lancet Respir Med. 2019;7(6):509–522. doi:10.1016/S2213-2600(19)30055-4

6. Lizzo JM, Goldin J, Cortes S. Pediatric Asthma. In: StatPearls. StatPearls Publishing; 2025. Accessed August 30, 2025. http://www.ncbi.nlm.nih.gov/books/NBK551631/

7. Sferrazza Papa GF, Pellegrino GM, Pellegrino R. Asthma and respiratory physiology: Putting lung function into perspective. Respirology. 2014;19(7):960–969. doi:10.1111/resp.12355

8. Shrine N, Izquierdo AG, Chen J, et al. Multi-ancestry genome-wide association analyses improve resolution of genes and pathways influencing lung function and chronic obstructive pulmonary disease risk. Nat Genet. 2023;55(3):410–422. doi:10.1038/s41588-023-01314-0

9. Wellcome Trust Case Control Consortium, The NSHD Respiratory Study Team, Repapi E, et al. Genome-wide association study identifies five loci associated with lung function. Nat Genet. 2010;42(1):36-44. doi:10.1038/ng.501

10. Global Initiative for Asthma (GINA). GINA Main Report 2024: Global Strategy for Asthma Management and Prevention. 2024. https://ginasthma.org/2024-report/

11. DeWan AT, Cahill ME, Cornejo-Sanchez DM, et al. Variants in JAZF1 are associated with asthma, type 2 diabetes, and height in the United Kingdom biobank population. Front Genet. 2023;14:1129389. doi:10.3389/fgene.2023.1129389

12. Mbatchou J, Barnard L, Backman J, et al. Computationally efficient whole-genome regression for quantitative and binary traits. Nat Genet. 2021;53(7):1097–1103. doi:10.1038/s41588-021-00870-7

13. Harpsøe MC, Basit S, Andersson M, et al. Body mass index and risk of autoimmune diseases: a study within the Danish National Birth Cohort. Int J Epidemiol. 2014;43(3):843–855. doi:10.1093/ije/dyu045

14. Purcell S, Neale B, Todd-Brown K, et al. PLINK: A Tool Set for Whole-Genome Association and Population-Based Linkage Analyses. Am J Hum Genet. 2007;81(3):559–575. doi:10.1086/519795

15. Chang CC, Chow CC, Tellier LC, Vattikuti S, Purcell SM, Lee JJ. Second-generation PLINK: rising to the challenge of larger and richer datasets. Gigascience. 2015;4(1):s13742-015-0047-0048. doi:10.1186/s13742-015-0047-8

16. Patterson N, Price AL, Reich D. Population Structure and Eigenanalysis. PLoS Genet. 2006;2(12):e190. doi:10.1371/journal.pgen.0020190

17. Bulik-Sullivan B, ReproGen Consortium, Psychiatric Genomics Consortium, et al. An atlas of genetic correlations across human diseases and traits. Nat Genet. 2015;47(11):1236–1241. doi:10.1038/ng.3406

18. Yang J, Benyamin B, McEvoy BP, et al. Common SNPs explain a large proportion of the heritability for human height. Nat Genet. 2010;42(7):565–569. doi:10.1038/ng.608

19. Yang J, Lee SH, Goddard ME, Visscher PM. GCTA: A Tool for Genome-wide Complex Trait Analysis. Am J Hum Genet. 2011;88(1):76–82. doi:10.1016/j.ajhg.2010.11.011

20. Lee SH, Wray NR, Goddard ME, Visscher PM. Estimating Missing Heritability for Disease from Genome-wide Association Studies. Am J Hum Genet. 2011;88(3):294–305. doi:10.1016/j.ajhg.2011.02.002

21. Watanabe K, Taskesen E, Van Bochoven A, Posthuma D. Functional mapping and annotation of genetic associations with FUMA. Nat Commun. 2017;8(1):1826. doi:10.1038/s41467-017-01261-5

22. Boughton AP, Welch RP, Flickinger M, et al. LocusZoom.js: interactive and embeddable visualization of genetic association study results. Marschall T, ed. Bioinformatics. 2021;37(18):3017–3018. doi:10.1093/bioinformatics/btab186

23. Wang G, Sarkar A, Carbonetto P, Stephens M. A Simple New Approach to Variable Selection in Regression, with Application to Genetic Fine Mapping. J R Stat Soc Ser B Stat Methodol. 2020;82(5):1273–1300. doi:10.1111/rssb.12388

24. Zou Y, Carbonetto P, Wang G, Stephens M. Fine-mapping from summary data with the “Sum of Single Effects” model. Epstein MP, ed. PLOS Genet. 2022;18(7):e1010299. doi:10.1371/journal.pgen.1010299

25. Zou Y, Carbonetto P, Xie D, Wang G, Stephens M. Fast and flexible joint fine-mapping of multiple traits via the Sum of Single Effects model. Genomics. Preprint posted online April 14, 2023. doi:10.1101/2023.04.14.536893

26. Tingley D, Yamamoto T, Hirose K, Keele L, Imai K. Mediation: R Package for Causal Mediation Analysis. J Stat Softw. 2014;59(5). doi:10.18637/jss.v059.i05

27. Lonsdale J, Thomas J, Salvatore M, et al. The Genotype-Tissue Expression (GTEx) project. Nat Genet. 2013;45(6):580–585. doi:10.1038/ng.2653

28. GTEx Consortium. The GTEx Consortium atlas of genetic regulatory effects across human tissues. Science. 2020;369(6509):1318–1330. doi:10.1126/science.aaz1776

29. Moffatt MF, Phil D, Gut IG, Strachan DP. A Large-Scale, Consortium-Based Genomewide Association Study of Asthma. N Engl J Med. Published online 2010.

30. Salinas YD, Wang Z, DeWan AT. Discovery and Mediation Analysis of Cross-Phenotype Associations Between Asthma and Body Mass Index in 12q13.2. Am J Epidemiol. 2021;190(1):85–94. doi:10.1093/aje/kwaa144

31. Zhang D, Zheng J. The Burden of Childhood Asthma by Age Group, 1990–2019: A Systematic Analysis of Global Burden of Disease 2019 Data. Front Pediatr. 2022;10:823399. doi:10.3389/fped.2022.823399

32. Van Rheenen W, Peyrot WJ, Schork AJ, Lee SH, Wray NR. Genetic correlations of polygenic disease traits: from theory to practice. Nat Rev Genet. 2019;20(10):567–581. doi:10.1038/s41576-019-0137-z

33. Feofanova EV, Brown MR, Alkis T, et al. Whole-Genome Sequencing Analysis of Human Metabolome in Multi-Ethnic Populations. Nat Commun. 2023;14(1):3111. doi:10.1038/s41467-023-38800-2

34. Tahir UA, Katz DH, Avila-Pachecho J, et al. Whole Genome Association Study of the Plasma Metabolome Identifies Metabolites Linked to Cardiometabolic Disease in Black Individuals. Nat Commun. 2022;13(1):4923. doi:10.1038/s41467-022-32275-3

35. Sun Y, Xu H, Ye K. GWAS and multi-omics integrative analysis reveal novel loci and their molecular mechanisms for circulating fatty acids. HGG Adv. 2025;6(4):100470. doi:10.1016/j.xhgg.2025.100470

36. Zhou F, Astle WJ, Butterworth AS, Asimit JL. Improved genetic discovery and fine-mapping resolution through multivariate latent factor analysis of high-dimensional traits. Cell Genomics. 2025;5(5):100847. doi:10.1016/j.xgen.2025.100847

37. Astle WJ, Elding H, Jiang T, et al. The Allelic Landscape of Human Blood Cell Trait Variation and Links to Common Complex Disease. Cell. 2016;167(5):1415–1429.e19. doi:10.1016/j.cell.2016.10.042

38. Chen MH, Raffield LM, Mousas A, et al. Trans-ethnic and Ancestry-Specific Blood-Cell Genetics in 746,667 Individuals from 5 Global Populations. Cell. 2020;182(5):1198–1213.e14. doi:10.1016/j.cell.2020.06.045

39. Krumsiek J, Suhre K, Evans AM, et al. Mining the unknown: a systems approach to metabolite identification combining genetic and metabolic information. PLoS Genet. 2012;8(10):e1003005. doi:10.1371/journal.pgen.1003005

40. Jia G, Zhong X, Im HK, et al. Discerning asthma endotypes through comorbidity mapping. Nat Commun. 2022;13(1):6712. doi:10.1038/s41467-022-33628-8

41. Zhou Y, Liang ZS, Jin Y, et al. Shared Genetic Architecture and Causal Relationship Between Asthma and Cardiovascular Diseases: A Large-Scale Cross-Trait Analysis. Front Genet. 2021;12:775591. doi:10.3389/fgene.2021.775591

42. Verma A, Huffman JE, Rodriguez A, et al. Diversity and scale: Genetic architecture of 2068 traits in the VA Million Veteran Program. Science. 2024;385(6706):eadj1182. doi:10.1126/science.adj1182

43. Sakaue S, Kanai M, Tanigawa Y, et al. A cross-population atlas of genetic associations for 220 human phenotypes. Nat Genet. 2021;53(10):1415–1424. doi:10.1038/s41588-021-00931-x

44. Vuckovic D, Bao EL, Akbari P, et al. The Polygenic and Monogenic Basis of Blood Traits and Diseases. Cell. 2020;182(5):1214–1231.e11. doi:10.1016/j.cell.2020.08.008

45. Kichaev G, Bhatia G, Loh PR, et al. Leveraging Polygenic Functional Enrichment to Improve GWAS Power. Am J Hum Genet. 2019;104(1):65–75. doi:10.1016/j.ajhg.2018.11.008

46. Graham SE, Clarke SL, Wu KHH, et al. The power of genetic diversity in genome-wide association studies of lipids. Nature. 2021;600(7890):675–679. doi:10.1038/s41586-021-04064-3

47. Jiang M, Hao X, Jiang Y, Li S, Wang C, Cheng S. Genetic and observational associations of lung function with gastrointestinal tract diseases: pleiotropic and mendelian randomization analysis. Respir Res. 2023;24(1):315. doi:10.1186/s12931-023-02621-0

48. Sinnott-Armstrong N, Tanigawa Y, Amar D, et al. Genetics of 35 blood and urine biomarkers in the UK Biobank. Nat Genet. 2021;53(2):185–194. doi:10.1038/s41588-020-00757-z

49. Zhu Z, Guo Y, Shi H, et al. Shared genetic and experimental links between obesity-related traits and asthma subtypes in UK Biobank. J Allergy Clin Immunol. 2020;145(2):537–549. doi:10.1016/j.jaci.2019.09.035

50. Tsuo K, Zhou W, Wang Y, et al. Multi-ancestry meta-analysis of asthma identifies novel associations and highlights the value of increased power and diversity. Cell Genomics. 2022;2(12):100212. doi:10.1016/j.xgen.2022.100212

51. Cosentino J, Behsaz B, Alipanahi B, et al. Inference of chronic obstructive pulmonary disease with deep learning on raw spirograms identifies new genetic loci and improves risk models. Nat Genet. 2023;55(5):787–795. doi:10.1038/s41588-023-01372-4

52. John C, Guyatt AL, Shrine N, et al. Genetic Associations and Architecture of Asthma-COPD Overlap. Chest. 2022;161(5):1155–1166. doi:10.1016/j.chest.2021.12.674

53. Yee SW, Buitrago D, Stecula A, et al. Deorphaning a solute carrier 22 family member, SLC22A15, through functional genomic studies. FASEB J Off Publ Fed Am Soc Exp Biol. 2020;34(12):15734-15752. doi:10.1096/fj.202001497R

54. Ohashi R, Tamai I, Yabuuchi H, et al. Na(+)-dependent carnitine transport by organic cation transporter (OCTN2): its pharmacological and toxicological relevance. J Pharmacol Exp Ther. 1999;291(2):778–784.

55. Wu X, Huang W, Prasad PD, et al. Functional characteristics and tissue distribution pattern of organic cation transporter 2 (OCTN2), an organic cation/carnitine transporter. J Pharmacol Exp Ther. 1999;290(3):1482–1492.

56. Wagner CA, Lükewille U, Kaltenbach S, et al. Functional and pharmacological characterization of human Na(+)-carnitine cotransporter hOCTN2. Am J Physiol Renal Physiol. 2000;279(3):F584–591. doi:10.1152/ajprenal.2000.279.3.F584

57. Maekawa S, Mori D, Nishiya T, et al. OCTN2VT, a splice variant of OCTN2, does not transport carnitine because of the retention in the endoplasmic reticulum caused by insertion of 24 amino acids in the first extracellular loop of OCTN2. Biochim Biophys Acta. 2007;1773(6):1000–1006. doi:10.1016/j.bbamcr.2007.04.005

58. Fujiya M, Inaba Y, Musch MW, Hu S, Kohgo Y, Chang EB. Cytokine regulation of OCTN2 expression and activity in small and large intestine. Inflamm Bowel Dis. 2011;17(4):907–916. doi:10.1002/ibd.21444

59. Sugiura T, Kato S, Shimizu T, et al. Functional Expression of Carnitine/Organic Cation Transporter OCTN1/SLC22A4 in Mouse Small Intestine and Liver. Drug Metab Dispos. 2010;38(10):1665–1672. doi:10.1124/dmd.110.032763

60. Pochini L, Scalise M, Galluccio M, Pani G, Siminovitch KA, Indiveri C. The human OCTN1 (SLC22A4) reconstituted in liposomes catalyzes acetylcholine transport which is defective in the mutant L503F associated to the Crohn’s disease. Biochim Biophys Acta BBA - Biomembr. 2012;1818(3):559–565. doi:10.1016/j.bbamem.2011.12.014

61. Gründemann D, Harlfinger S, Golz S, et al. Discovery of the ergothioneine transporter. Proc Natl Acad Sci U S A. 2005;102(14):5256–5261. doi:10.1073/pnas.0408624102

62. Guryanova OA, Drazba JA, Frolova EI, Chumakov PM. Actin Cytoskeleton Remodeling by the Alternatively Spliced Isoform of PDLIM4/RIL Protein. J Biol Chem. 2011;286(30):26849–26859. doi:10.1074/jbc.M111.241554

63. Qi H, Zhu H, Lou M, et al. Interferon regulatory factor 1 transactivates expression of human DNA polymerase η in response to carcinogen N-methyl-N’-nitro-N-nitrosoguanidine. J Biol Chem. 2012;287(16):12622–12633. doi:10.1074/jbc.M111.313429

64. Oshima S, Nakamura T, Namiki S, et al. Interferon regulatory factor 1 (IRF-1) and IRF-2 distinctively up-regulate gene expression and production of interleukin-7 in human intestinal epithelial cells. Mol Cell Biol. 2004;24(14):6298–6310. doi:10.1128/MCB.24.14.6298-6310.2004

65. Dornan D, Eckert M, Wallace M, et al. Interferon regulatory factor 1 binding to p300 stimulates DNA-dependent acetylation of p53. Mol Cell Biol. 2004;24(22):10083–10098. doi:10.1128/MCB.24.22.10083-10098.2004

66. Su ZZ, Sarkar D, Emdad L, Barral PM, Fisher PB. Central role of interferon regulatory factor-1 (IRF-1) in controlling retinoic acid inducible gene-I (RIG-I) expression. J Cell Physiol. 2007;213(2):502–510. doi:10.1002/jcp.21128

67. Park J, Kim K, Lee EJ, et al. Elevated level of SUMOylated IRF-1 in tumor cells interferes with IRF-1-mediated apoptosis. Proc Natl Acad Sci U S A. 2007;104(43):17028–17033. doi:10.1073/pnas.0609852104

68. Bowie ML, Ibarra C, Seewalt VL. IRF-1 promotes apoptosis in p53-damaged basal-type human mammary epithelial cells: a model for early basal-type mammary carcinogenesis. Adv Exp Med Biol. 2008;617:367–374. doi:10.1007/978-0-387-69080-3_35

69. Huang Y, Walstrom A, Zhang L, et al. Type I interferons and interferon regulatory factors regulate TNF-related apoptosis-inducing ligand (TRAIL) in HIV-1-infected macrophages. PloS One. 2009;4(4):e5397. doi:10.1371/journal.pone.0005397

70. Gao J, Senthil M, Ren B, et al. IRF-1 transcriptionally upregulates PUMA, which mediates the mitochondrial apoptotic pathway in IRF-1-induced apoptosis in cancer cells. Cell Death Differ. 2010;17(4):699–709. doi:10.1038/cdd.2009.156

71. Weng X, Zheng M, Liu Y, Lou G. The role of Bach2 in regulating CD8 + T cell development and function. Cell Commun Signal. 2024;22(1):169. doi:10.1186/s12964-024-01551-8

72. Liu G, Liu F. Bach2: A Key Regulator in Th2-Related Immune Cells and Th2 Immune Response. Xu H, ed. J Immunol Res. 2022;2022:1–10. doi:10.1155/2022/2814510

73. Fry A, Littlejohns TJ, Sudlow C, et al. Comparison of Sociodemographic and Health-Related Characteristics of UK Biobank Participants With Those of the General Population. Am J Epidemiol. 2017;186(9):1026–1034. doi:10.1093/aje/kwx246

