## Supplemental Figure and Table legends for "Shared genetic factors between lung function and asthma by age at onset"

**Supplementary figures legends**

Supplementary figure 1. Venn diagrams for overlapping variants (A) and pathways (B) between asthma age-at-onset groups and lung function. The total number of mapped variants/pathways for each trait is provided below the name.

Supplementary figure 2. An example of region selection flowchart between asthma LT20 and lung function.

Supplementary figure 3. Example LocusZoom plot on chromosome 17: 38103000-38410000 (hg19) between asthma LT20 and lung function. The top variant in discovery is highlighted as the lead variant with summary statistics displayed from both discovery and replication data in each trait. (A) asthma LT20 discovery (B) asthma LT20 replication (C) lung function discovery (D) lung function replication.

Supplementary figure 4. Univariate and multivariate fine mapping on chromosome 5. A. SuSiE for asthma LT20. B. SuSiE for asthma 20-40. C. SuSiE for asthma GT40. D. SuSiE for lung function. E. mvSuSiE PIP plot. Purity: minimum absolute correlation in the CS. PIP: posterior inclusion probability. L: the CS identified. C: number of SNVs in the CS.

Supplementary figure 5. Univariate and multivariate fine mapping on chromosome 6. A. SuSiE for asthma LT20. B. SuSiE for asthma 20-40. C. SuSiE for lung function. D. mvSuSiE PIP plot. Purity: minimum absolute correlation in the CS. PIP: posterior inclusion probability. L: the CS identified. C: number of SNVs in the CS.

Supplementary figure 6. Univariate and multivariate fine mapping on chromosome 12. A. SuSiE for asthma LT20. B. SuSiE for asthma 20-40. C. SuSiE for lung function. D. mvSuSiE PIP plot. Purity: minimum absolute correlation in the CS. PIP: posterior inclusion probability. L: the CS identified. C: number of SNVs in the CS.

Supplementary figure 7. Univariate and multivariate fine mapping on chromosome 17. A. SuSiE for asthma LT20. B. SuSiE for lung function. C. mvSuSiE PIP plot. Purity: minimum absolute correlation in the CS. PIP: posterior inclusion probability. L: the CS identified. C: number of SNVs in the CS.

Supplementary figure 8. GWAS for asthma LT20. A. Manhattan plot. B. Q-Q plot. Analysis was performed in REGENIE and adjusted for age at recruitment, genetic sex, and top 10 PCs. λ: genomic inflation factor.

Supplementary figure 9. GWAS for asthma 20-40. A. Manhattan plot. B. Q-Q plot. Analysis was performed in REGENIE and adjusted for age at recruitment, genetic sex, and top 10 PCs. λ: genomic inflation factor.

Supplementary figure 10. GWAS for asthma GT40. A. Manhattan plot. B. Q-Q plot. Analysis was performed in REGENIE and adjusted for age at recruitment, genetic sex, and top 10 PCs. λ: genomic inflation factor.

Supplementary figure 11. GWAS for lung function. A. Manhattan plot. B. Q-Q plot. Analysis was performed in REGENIE and adjusted for age at recruitment, genetic sex, top 10 PCs, and baseline smoking status. λ: genomic inflation factor.

Supplementary figure 12. GWAS for COPD. A. Manhattan plot. B. Q-Q plot. Analysis was performed in REGENIE and adjusted for age at recruitment, genetic sex, top 10 PCs, and baseline smoking status. λ: genomic inflation factor.

**Supplementary tables**

Supplementary table 1a. Asthma grouping using LDSC -- step 1

Supplementary table 1b. Asthma grouping using LDSC -- step 2

Supplementary table 1c. Asthma grouping using LDSC -- step 3

Supplementary table 1d. Details for heritability estimation using GCTA

Supplementary table 2a. Replication summary statistics for overlapping variants (asthma LT20, 20-40, GT40, and lung function) in discovery data

Supplementary table 2b. Replication summary statistics for overlapping variants (asthma LT20, 20-40, and lung function) in discovery data

Supplementary table 2c. Replication summary statistics for overlapping variants (asthma LT20, GT40, and lung function) in discovery data

Supplementary table 2d. Replication summary statistics for overlapping variants (asthma LT20 and lung function) in discovery data

Supplementary table 2e. Replication summary statistics for overlapping variants (asthma GT40 and lung function) in discovery data

Supplementary table 3. List of overlapping genes between asthma subtypes and lung function

Supplementary table 4. Lead variant summary statistics in selected regions

Supplementary table 5a. Chromosome 5 region fine mapping

Supplementary table 5b. Chromosome 6 region fine mapping

Supplementary table 5c. Chromosome 12 region fine mapping

Supplementary table 5d. Chromosome 17 region fine mapping

Supplementary table 6a. List of *cis* eQTL genes

Supplementary table 6b. List of *cis* sQTL genes
