## Supplemental Figures for "Shared genetic factors between lung function and asthma by age at onset"

### Slide 1
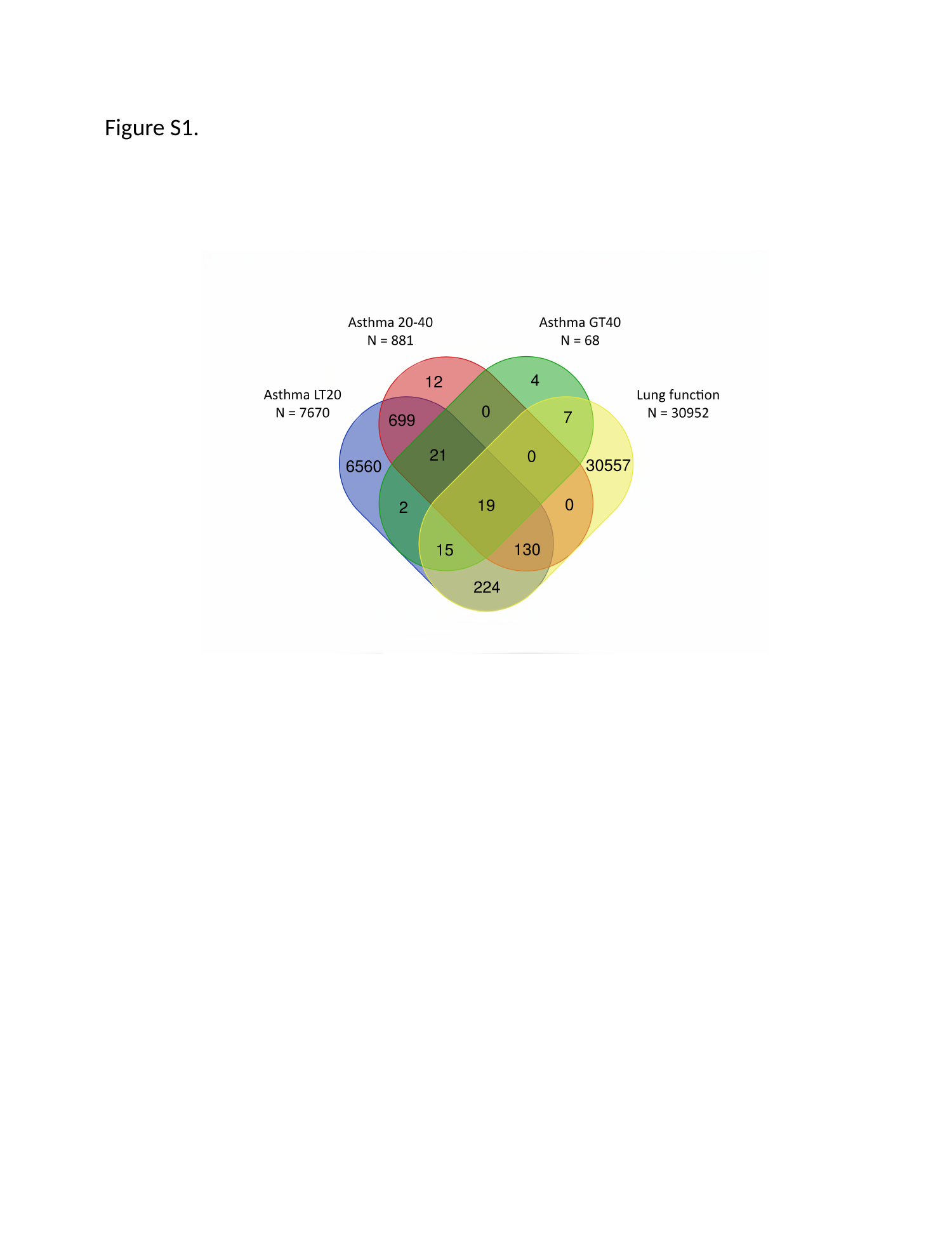

Figure S1.

### Slide 2
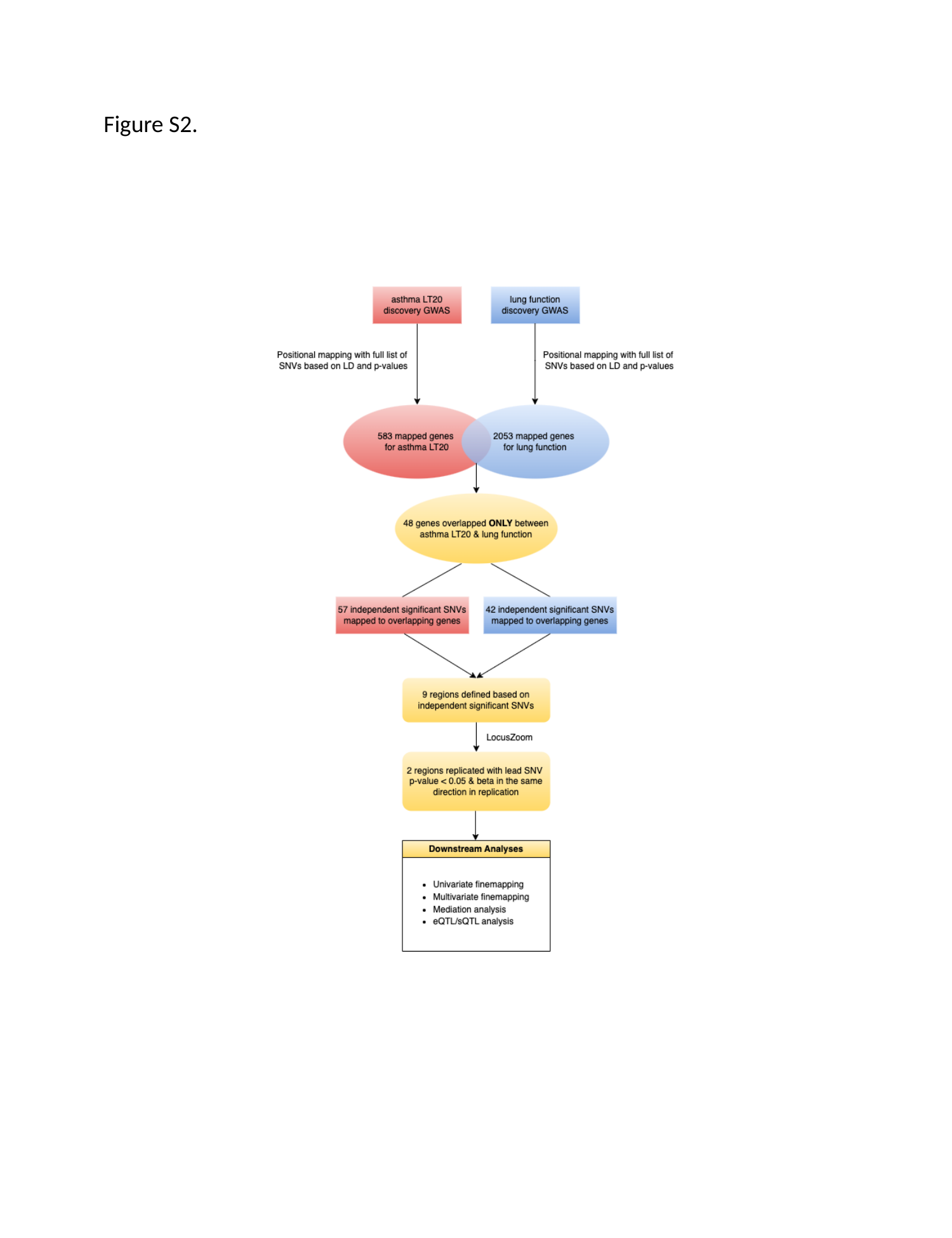

Figure S2.

### Slide 3
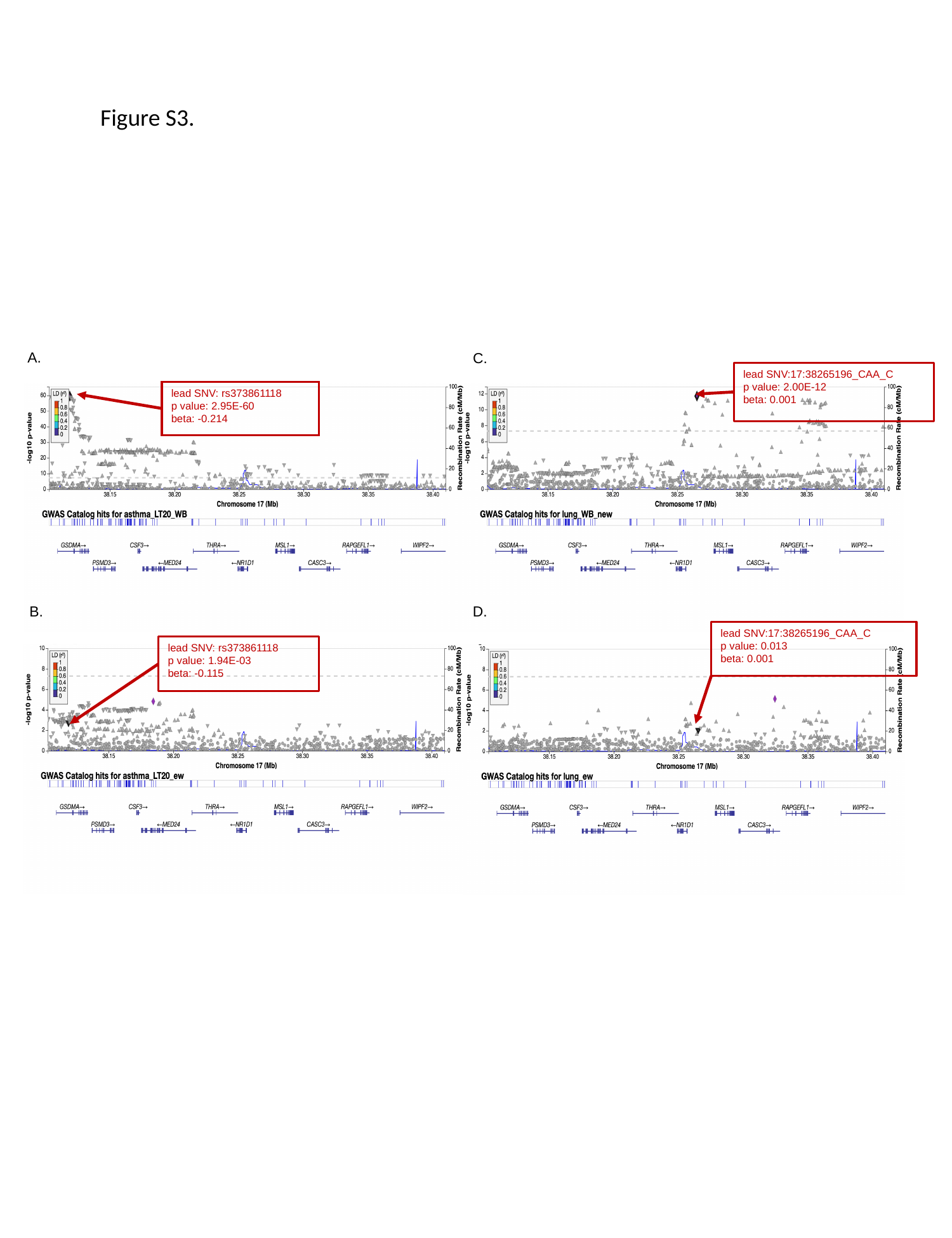

Figure S3.
A.
C.
lead SNV:17:38265196_CAA_C
p value: 2.00E-12
beta: 0.001
lead SNV: rs373861118
p value: 2.95E-60
beta: -0.214
D.
B.
lead SNV:17:38265196_CAA_C
p value: 0.013
beta: 0.001
lead SNV: rs373861118
p value: 1.94E-03
beta: -0.115

### Slide 4
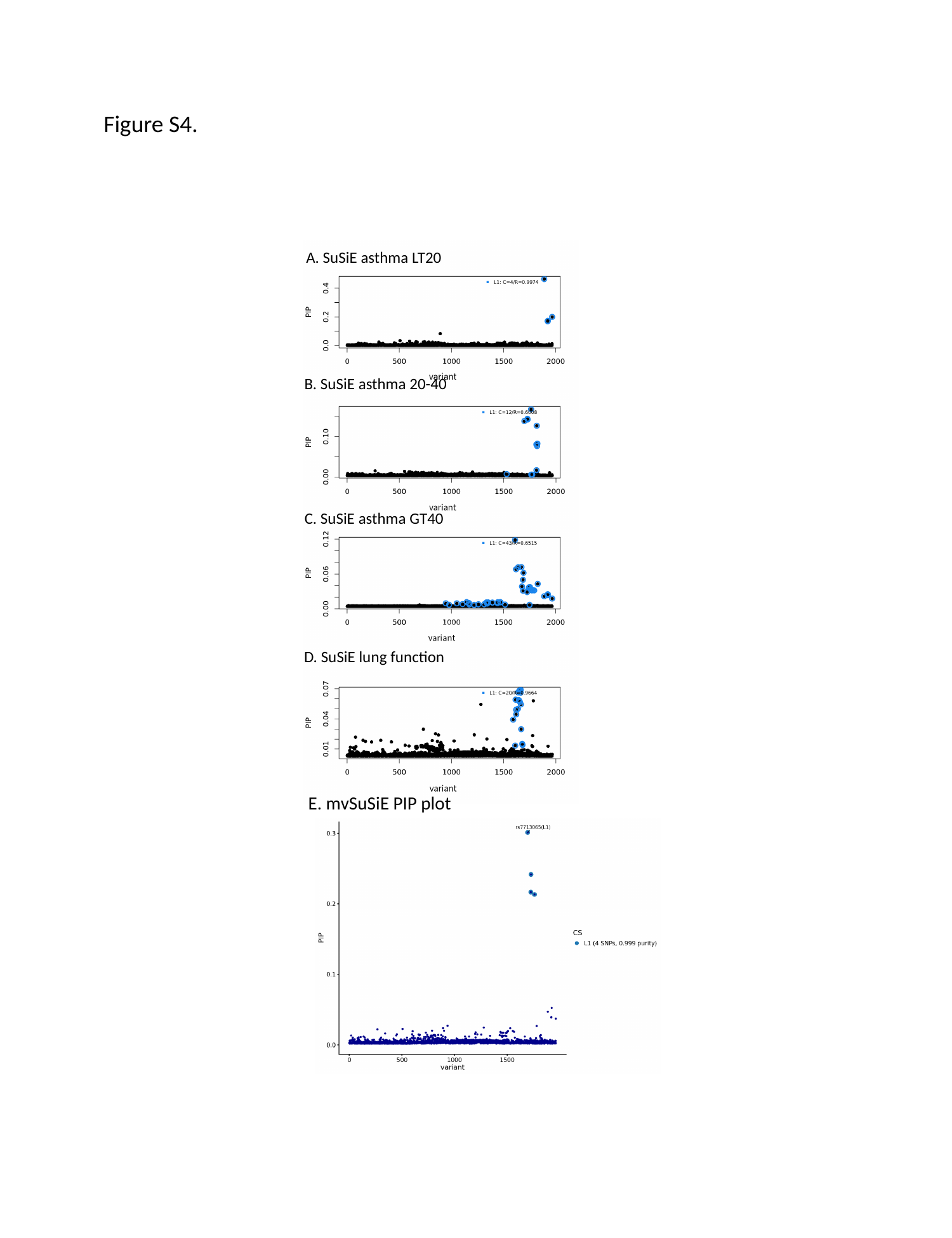

Figure S4.
A. SuSiE asthma LT20
B. SuSiE asthma 20-40
C. SuSiE asthma GT40
D. SuSiE lung function
E. mvSuSiE PIP plot

### Slide 5
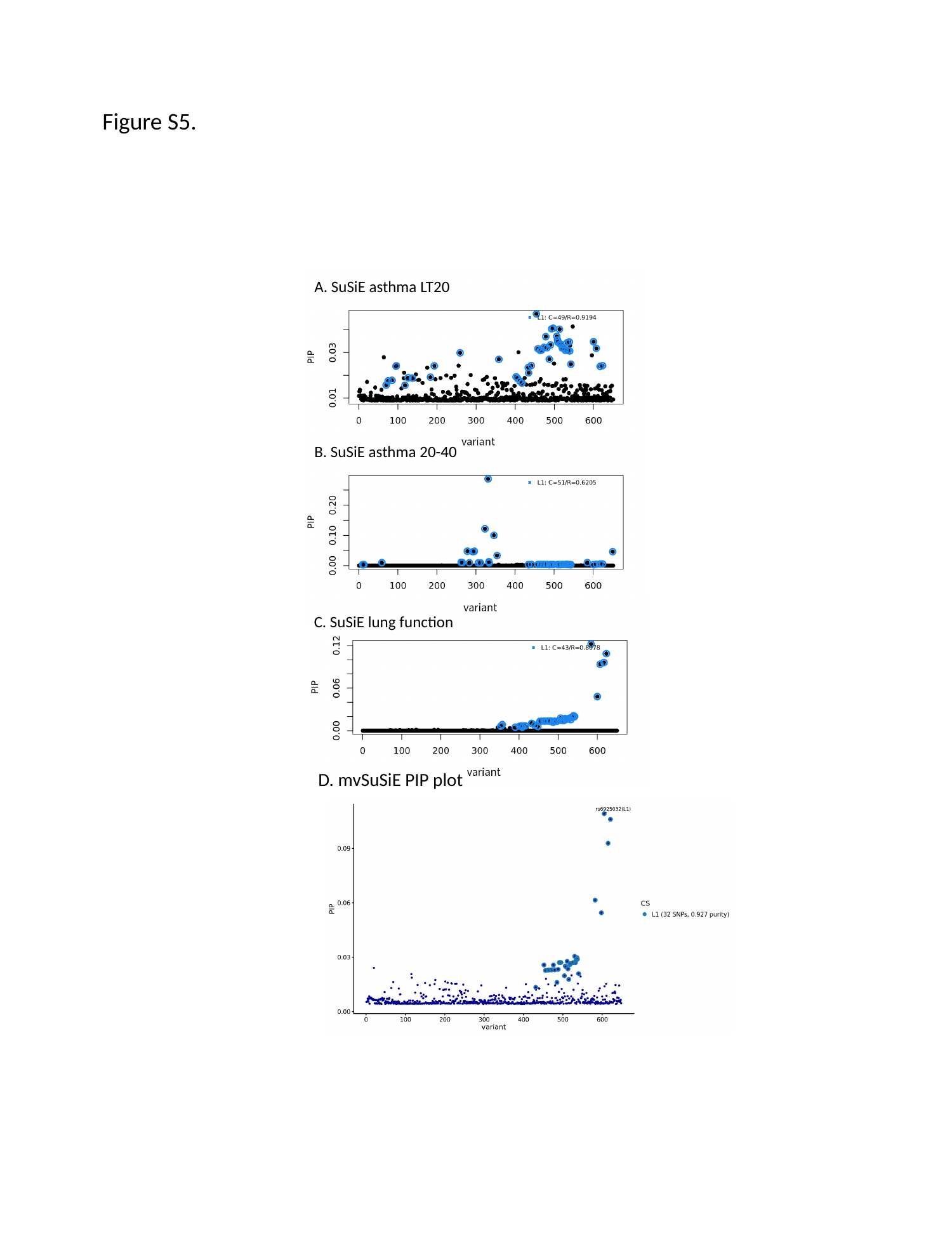

Figure S5.
A. SuSiE asthma LT20
B. SuSiE asthma 20-40
C. SuSiE lung function
D. mvSuSiE PIP plot

### Slide 6
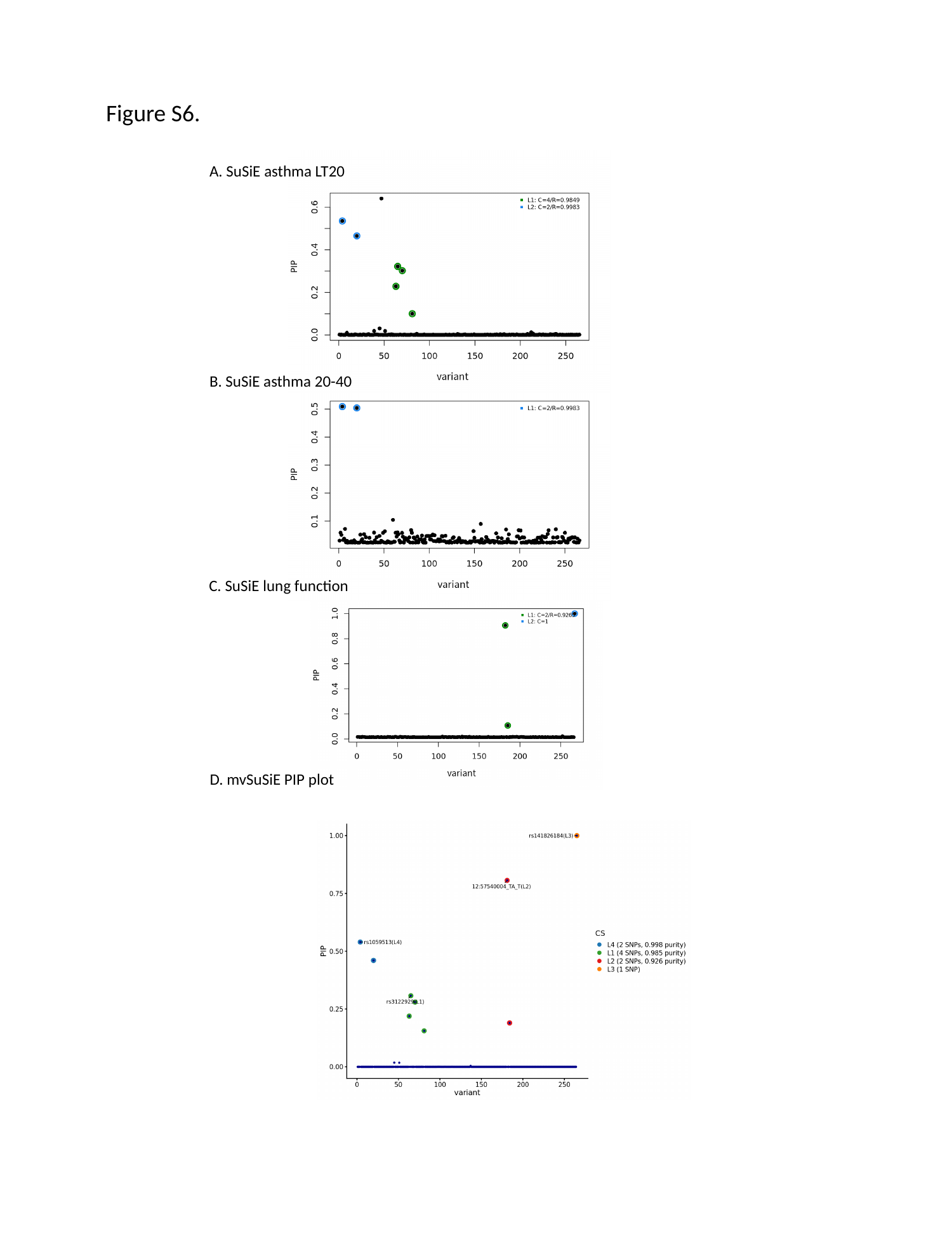

Figure S6.
A. SuSiE asthma LT20
B. SuSiE asthma 20-40
C. SuSiE lung function
D. mvSuSiE PIP plot

### Slide 7
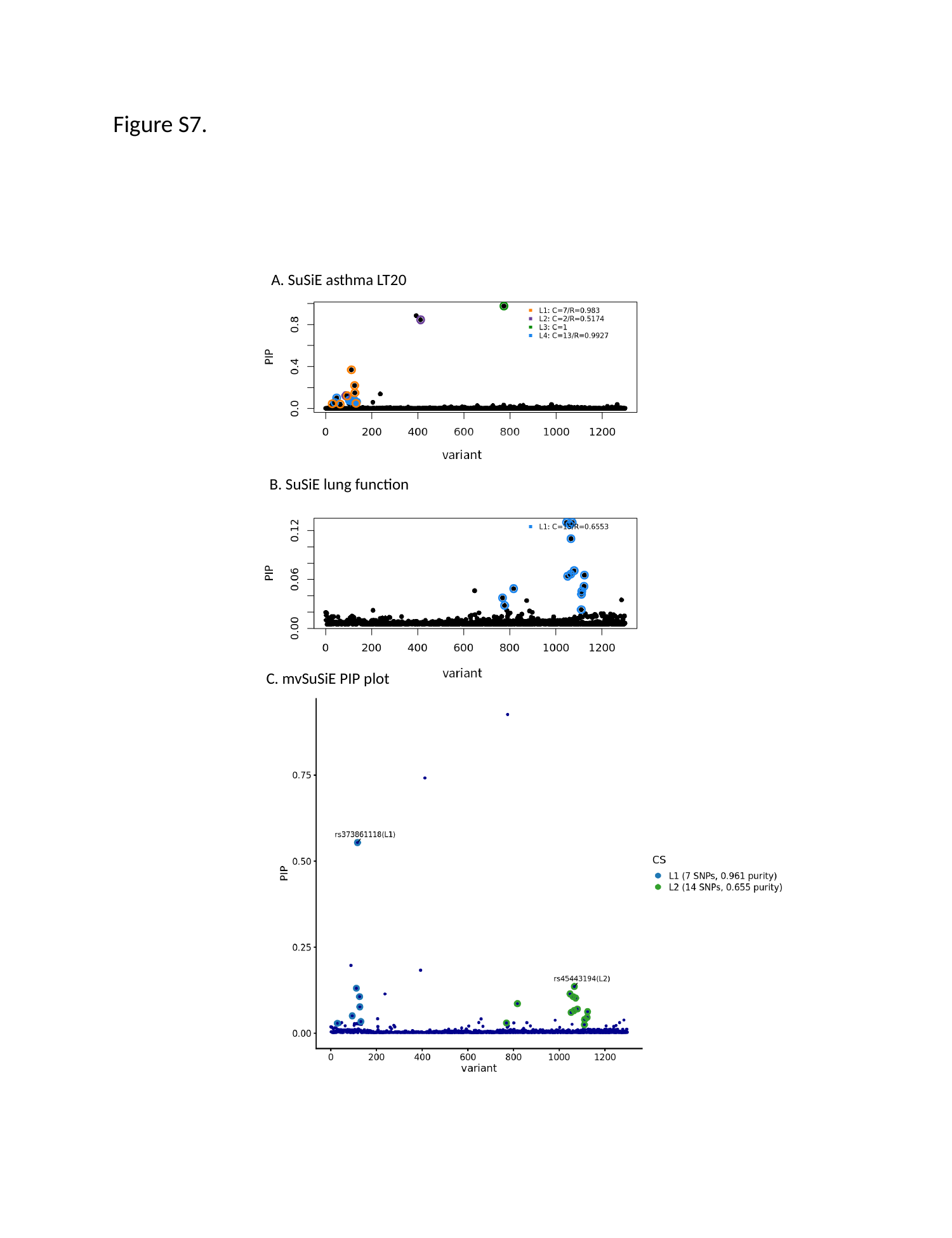

Figure S7.
A. SuSiE asthma LT20
B. SuSiE lung function
C. mvSuSiE PIP plot

### Slide 8
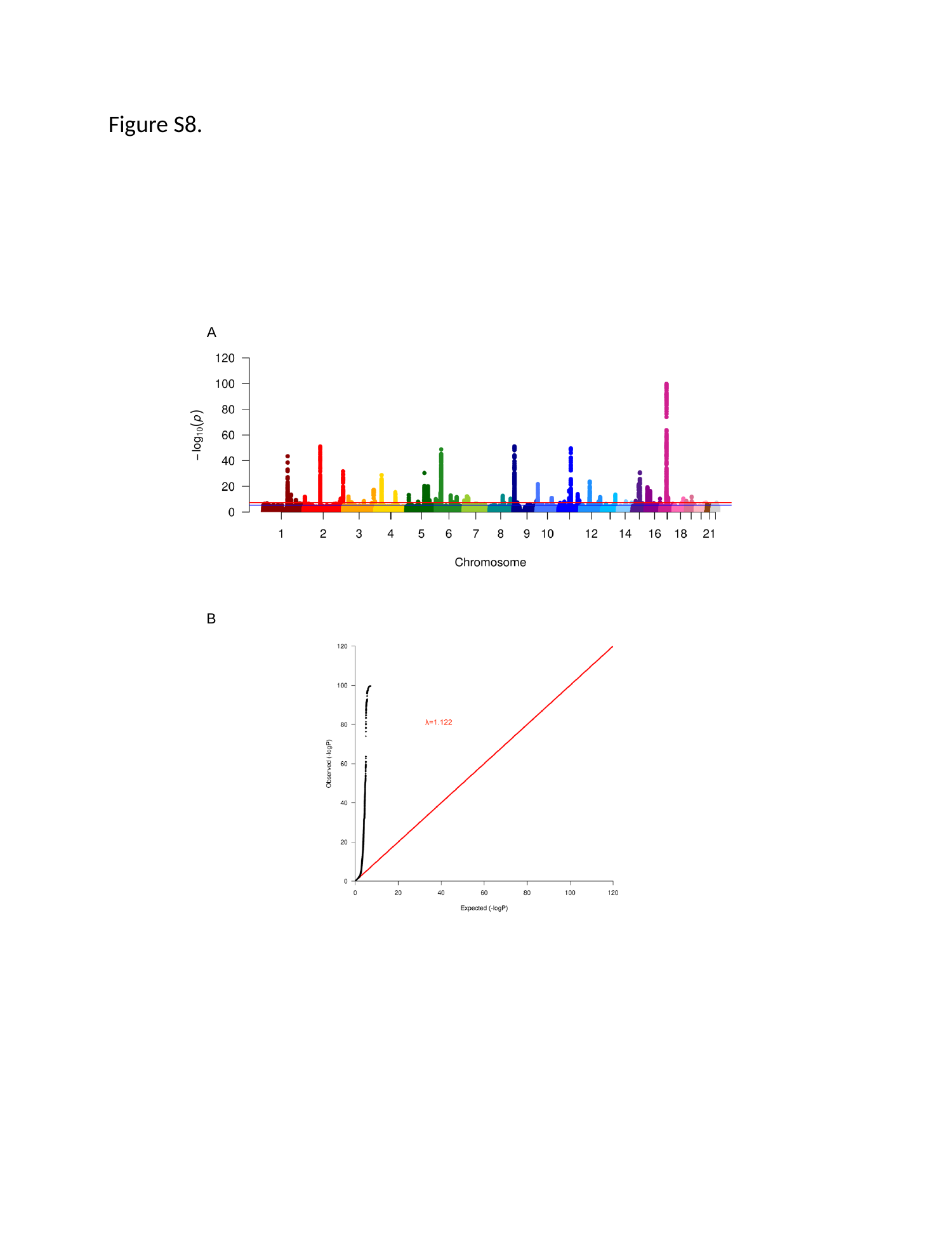

Figure S8.
A
B

### Slide 9
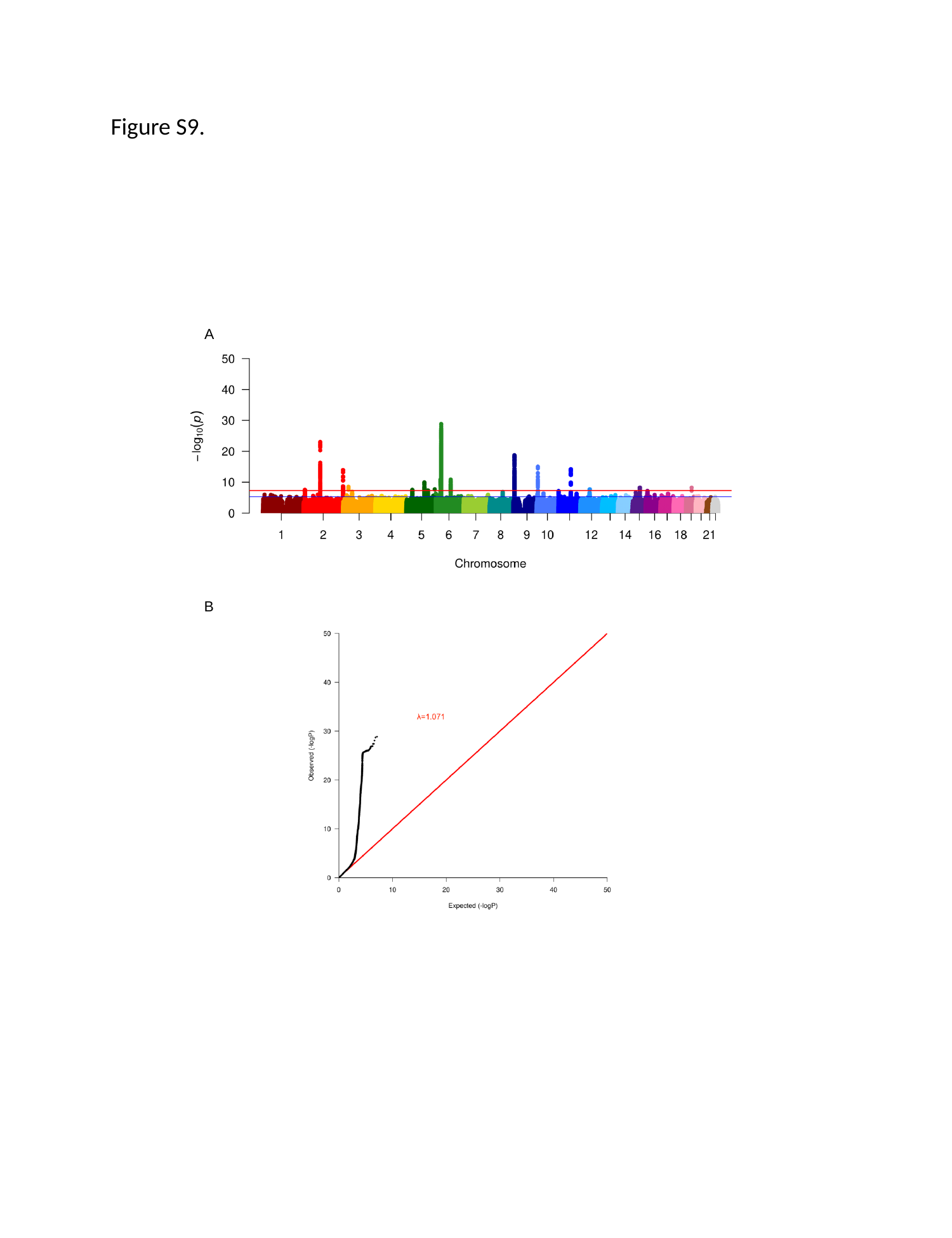

Figure S9.
A
B

### Slide 10
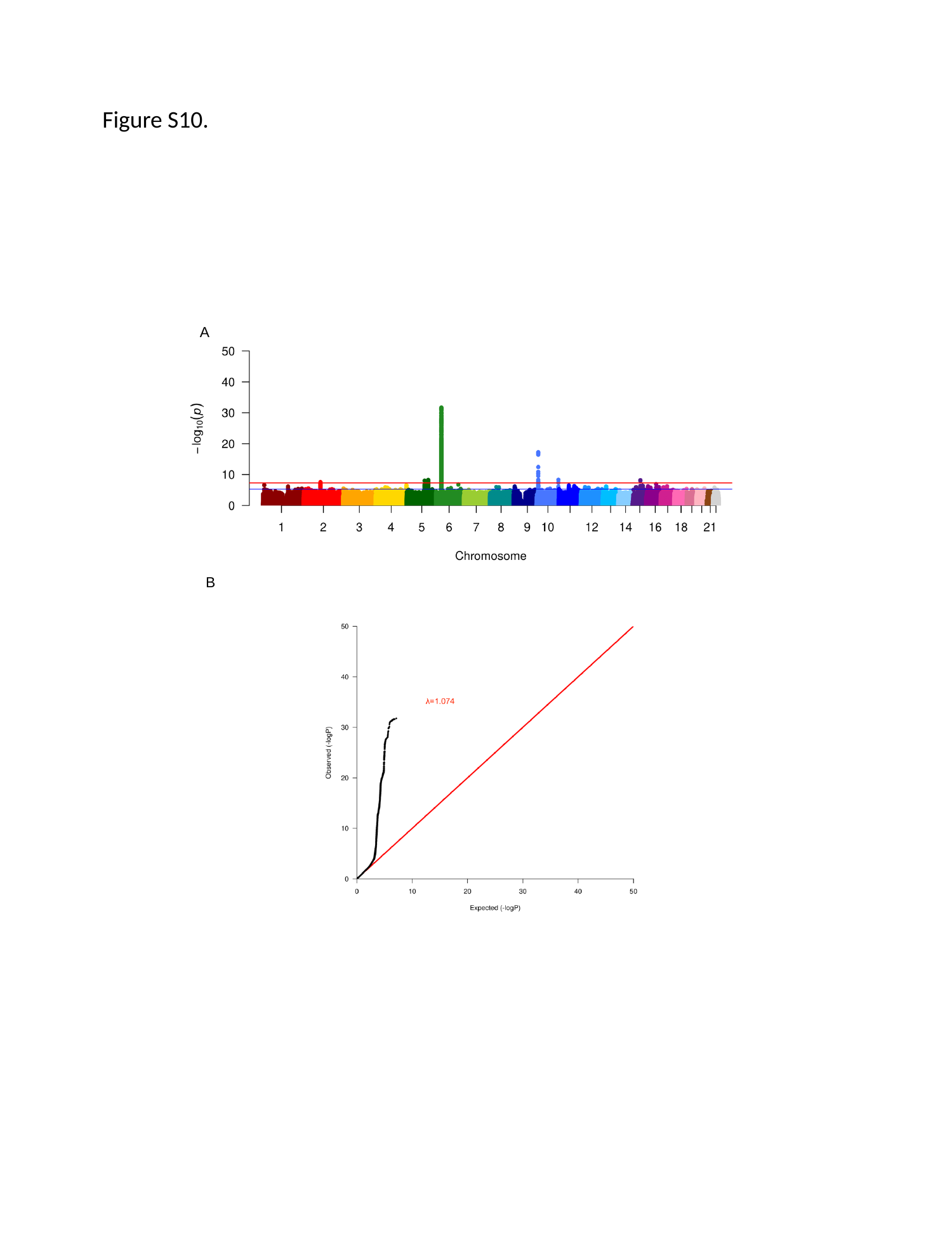

Figure S10.
A
B

### Slide 11
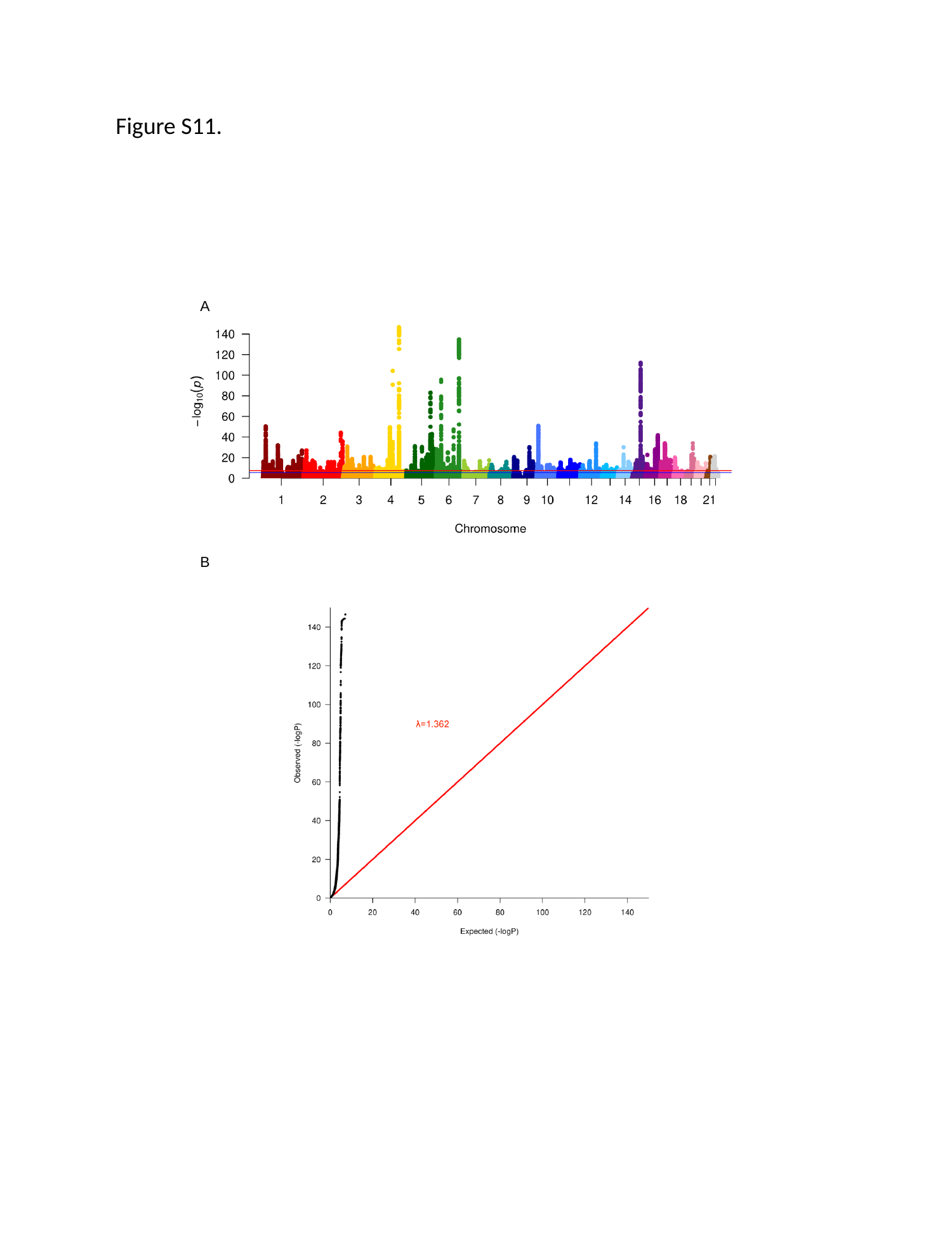

Figure S11.
A
B

### Slide 12
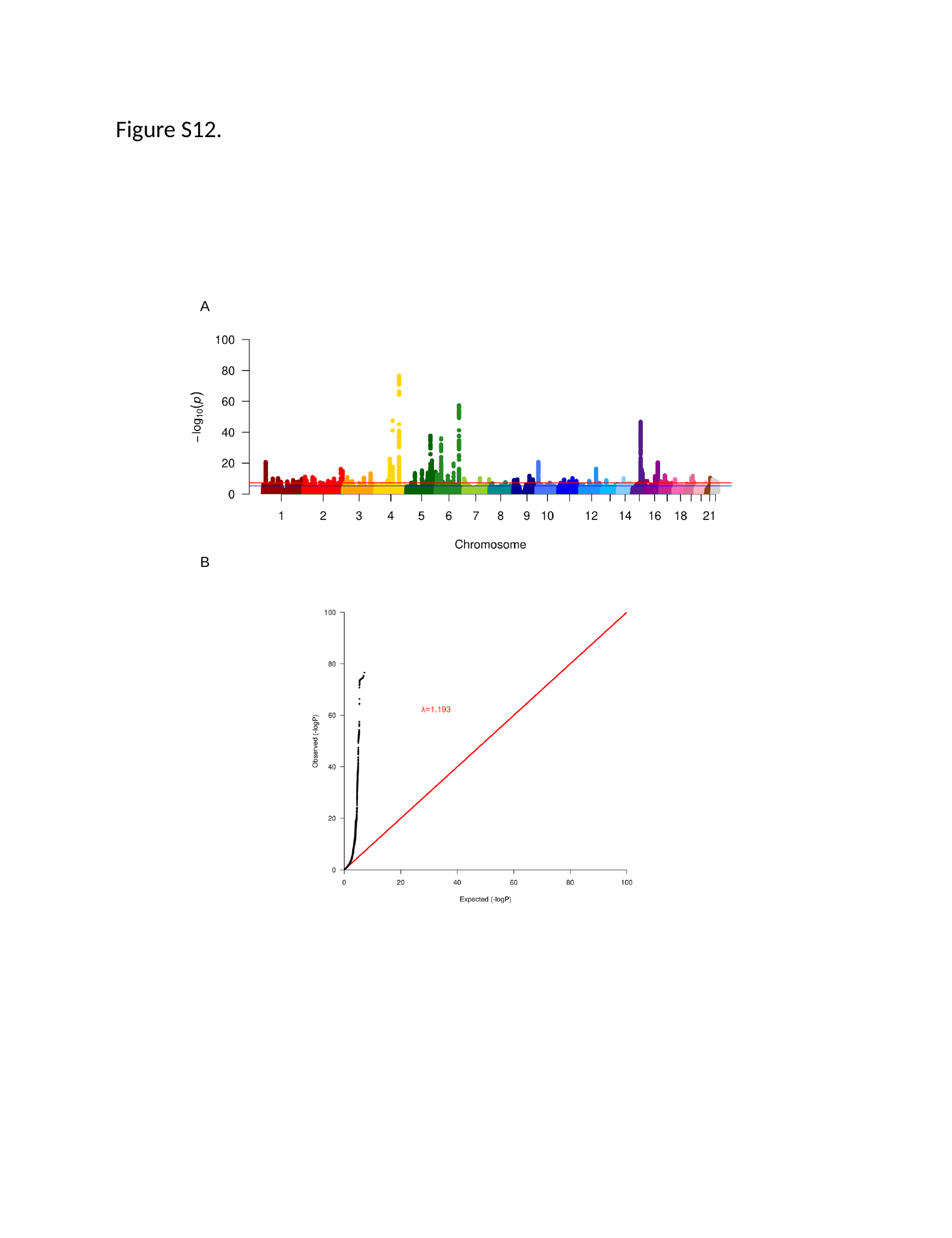

Figure S12.
A
B
